# Second round of the interlaboratory comparison (ILC) exercise of SARS-CoV-2 molecular detection assays being used by 45 veterinary diagnostic laboratories in the US

**DOI:** 10.1101/2022.04.08.22273621

**Authors:** Kaiping Deng, Steffen Uhlig, Laura B. Goodman, Hon S. Ip, Mary Lea Killian, Sarah M. Nemser, Jodie Ulaszek, Shannon Kiener, Matthew Kmet, Kirstin Frost, Karina Hettwer, Bertrand Colson, Kapil Nichani, Anja Schlierf, Andriy Tkachenko, Mothomang Mlalazi-Oyinloye, Andrew Scott, Ravinder Reddy, Gregory H. Tyson

## Abstract

The coronavirus disease 2019 (COVID-19) pandemic presents a continued public health challenge across the world. Veterinary diagnostic laboratories in the U.S. use real-time reverse transcriptase PCR (RT-PCR) for animal testing, and many are certified for testing human samples, so ensuring laboratories have sensitive and specific SARS-CoV-2 testing methods is a critical component of the pandemic response. In 2020, the FDA Veterinary Laboratory Investigation and Response Network (Vet-LIRN) led the first round of an Inter-Laboratory Comparison (ILC) Exercise to help laboratories evaluate their existing real-time RT-PCR methods for detecting SARS-CoV-2. The ILC1 results indicated that all participating laboratories were able to detect the viral RNA spiked in buffer and PrimeStore molecular transport medium (MTM). The current ILC (ILC2) aimed to extend ILC1 by evaluating analytical sensitivity and specificity of the methods used by participating laboratories to detect three SARS-CoV-2 variants (B.1, B.1.1.7 (Alpha) and B.1.351 (Beta)). ILC2 samples were prepared with RNA at levels between 10 to 10,000 copies per 50 μL MTM. Fifty-seven sets of results from 45 laboratories were qualitatively and quantitatively analyzed according to the principles of ISO 16140-2:2016. The results showed that over 95% of analysts detected the SARS-CoV-2 RNA in MTM at 500 copies or higher for all three variants. In addition, 81% and 92% of the analysts achieved a Level of Detection (LOD95_eff. vol._) below 20 copies in the assays with nucleocapsid markers N1 and N2, respectively. The analytical specificity of the evaluated methods was over 99%. The study allowed participating laboratories to assess their current method performance, identify possible limitations, and recognize method strengths as part of a continuous learning environment to support the critical need for reliable diagnosis of COVID-19 in potentially infected animals and humans.

---

Emergence of Severe Acute Respiratory Syndrome Coronavirus (SARS-CoV) in 2003^1^ and the Middle Eastern Respiratory Syndrome Coronavirus (MERS-CoV) in 2012^2^ showed the zoonotic potential of animal coronaviruses. SARS-CoV-2-transmission has been documented among animals, from humans to animals, and from animals to humans in the COVID-19 pandemic^1,3-7^. SARS-CoV-2 has been detected in wild and domestic animals around the world^8-17^. A recent U.S. animal surveillance study indicated high prevalence of SARS-CoV-2 among domestic and wild free-roaming animals tested on mink farms^18^. Conducting animal surveillance and routine testing with a sensitive and specific SARS-CoV-2 detection method is important in outbreak response and prevention. A joint statement from United Nations Food and Agriculture Organisation (FAO)/World Organization for Animal Health (OIE)/World Health Organisation (WHO) also noted the need to promote the monitoring of wildlife known to be potentially susceptible to SARS-CoV-2 and reporting of confirmed animal cases of SARS-CoV-2 to OIE, with these actions requiring a sensitive and accurate SARS-CoV-2 test.

In response to the spread of SARS-CoV-2 in animals, veterinary diagnostic laboratories receive animal specimens for its detection. Many veterinary diagnostic laboratories also have Clinical Laboratory Improvement Amendments (CLIA) certification and test human samples, meaning they play a critical One Health role in assessing the impact of COVID-19 on both humans and animals. Among the available diagnostic tests, detection of the viral RNA with real-time reverse transcriptase polymerase chain reaction (RT-PCR) is the most widely used, sensitive, and specific diagnostic method for COVID-19.

The U.S. FDA’s Veterinary Laboratory Investigation and Response Network (Vet-LIRN) is a network of veterinary diagnostic laboratories that investigates potential animal food or drug related issues^19^. In August 2020, an Inter-Laboratory Comparison Exercise Round 1 (ILC1) was collaboratively conducted by FDA and other organizations to qualitatively and quantitatively evaluate the SARS-CoV-2 real-time RT-PCR detection methods used by veterinary diagnostic laboratories^20^. The results indicated that the ILC1 participants effectively detected SARS-CoV-2 RNA in MTM with their methods routinely used for testing clinical specimens. Two-thirds of the laboratories achieved nearly the theoretical optimum Level of Detection (LOD) of three copies^20^. However, the viral RNA spiking levels of ILC1 were not low enough to evaluate the method analytical sensitivity, specifically the LOD for each individual participant. Hence, this second round of ILC (ILC2) was designed to provide more challenging samples from which marginal detection results could be generated for statistical analysis.

New lineages of SARS-CoV-2 were reported and quickly became dominant variants in different parts of the world since late 2020^21^ (https://covariants.org/), including the Alpha variant (B.1.1.7) and the Beta variant (B.1.351)^22,23^. Additional variants of concern such as the Delta variant (B.1.617) and Omicron variant (B.1.1.529) subsequently emerged. These emerging variants carry numerous mutations throughout the viral genome, including on the spike (*S*), envelope (*E*), nucleocapsid (*N*) and/or *ORF1ab* genes. Most participants used assays detecting the *N* gene in their routine testing, specifically markers N1 and N2, whereas some laboratories used other gene markers such as *ORF1ab, S* or *E* genes. In ILC2, the variants designated as Alpha (B.1.1.7) and Beta (B.1.351) were used in addition to U.S. B.1, which was the most prevalent lineage in the U.S. at the end of 2020 and the beginning of 2021. The ILC2 samples were shipped to laboratories in June of 2021. Detection of these variants with different markers used by participants was further studied in ILC2.

ILC2 was collaboratively conducted by the following: (i) the FDA’s Center for Veterinary Medicine’s Vet-LIRN, (ii) the Moffett Proficiency Testing (PT) Laboratory located at the Institute for Food Safety and Health at the Illinois Institute of Technology (IIT/IFSH) and the FDA Division of Food Processing Science and Technology, (iii) QuoData Quality and Statistics GmbH in Germany, (iv) Cornell University, (v) the U.S. Geological Survey (USGS) National Wildlife Center, (vi) USDA’s National Veterinary Services Laboratories (NVSL), National Animal Health Laboratory Network (NAHLN), (vii) the Integrated Consortium of Laboratory Networks (ICLN), and (viii) 45 participating laboratories. The study was a continuation of the previous ILC1 with the following objectives: (1) to evaluate analytical sensitivity (i.e., LOD) of the methods routinely used by participating laboratories to detect SARS-CoV-2 variant (B.1) RNA; (2) to evaluate the ability of the participants’ methods to detect SARS-CoV-2 variants Alpha (B.1.1.7) and Beta (B.1.351); and (3) to evaluate the methods’ specificity by investigating cross-reactivity with a non-SARS-CoV-2 animal coronavirus, Feline Infectious Peritonitis virus (FIPV) RNA. The goal was to allow participating laboratories to assess their method performance, including strengths and limitations, to support reliable diagnosis of COVID-19 in potentially infected animals and humans.

## Materials and methods

### Determination of SARS-CoV-2 RNA concentration

Three synthetic SARS-CoV-2 genomic RNA products were purchased from Twist Bioscience for the ILC2 study: B.1 (Twist Control #10), Alpha (B.1.1.7) (Twist Control #15) and Beta (B.1.351) (Twist Control #16). These ssRNA controls are manufactured by *in vitro* transcription from six non-overlapping 5 kb synthetic gene fragments. According to the manufacturer, the synthetic RNAs cover 99.9% of the bases of the viral genomes that were predominant in the U.S., including the USA/CA-PC101P/2020 (B.1), United Kingdom [Alpha (B.1.1.7)] and South Africa [Beta (B.1.351)] variants (GISAID names: USA/CA-PC101P/2020, England/205041766/2020 and South Africa/KRISP-EC-K005299/2020), respectively. Droplet digital reverse-transcription PCR (RT-ddPCR)-based quantification of these controls was performed by the Cornell University Genomics Facility using the QX200 instrument (Bio-Rad). The CDC N1 primers and probe (IDT) were used for this analysis with the 1-Step RT-ddPCR advanced kit for probes (Bio-Rad), on duplicate serial dilutions of the templates. The concentrations of the original Twist B.1, Alpha (B.1.1.7), and Beta (B.1.351) controls were determined by RT-ddPCR as 150,000, 345,000, and 300,000 copies/μL, respectively. Serial dilutions of the controls were then made in Nucleic Acid Dilution Solution (NADS) from the VetMAX™ Xeno™ Internal Positive Control DNA kit (Applied Biosystems) to levels of 2 × 10^5^ to 2 copies per μL in ten-fold dilutions.

### Feline Infectious Peritonitis virus (FIPV) RNA preparation

A cryopreserved suspension of the culture-adapted Black strain of FIPV was provided by Dr. Gary Whittaker at Cornell University. The culture was grown in Fcwf-4 CU cells as previously described^24^. RNA was extracted and purified using the MagMAX™ Viral/Pathogen kit (Thermo Fisher). Quantification by RT-ddPCR was performed as described above but with the P009 and P010 primers and P9/ P10 probe^25^, that targets the N gene of FIPV.

### ILC2 sample stability and homogeneity studies

Acceptable homogeneity, stability, and targeted spike levels were verified in three studies. In the first study (Study-1), samples were analyzed by two analysts in two trials on days 0, 7, 14, and 21 of storage at -80 °C. Eighteen samples (S1 to S18) for each set were prepared. S1-S16 were prepared by adding B.1 RNA to PrimeStore Molecular Transport Medium (MTM, Longhorn Vaccines & Diagnostics, LLC) at levels of 0, 10, 20, 50, 100, or 1,000 copies per 50 μL. S17 and S18 were prepared by adding Alpha (B.1.1.7) and Beta (B.1.351) to MTM at 1,000 copies per 50 μL. The RNA in Study-1 samples were isolated using Qiagen RNeasy Mini kit (Qiagen) and subsequently analyzed with AgPath-ID™ One-Step RT-PCR kit using specific primers (i.e., N1 and N2 targeting two regions of the viral nucleocapsid gene) and probes for the virus N gene (IDT), according to the CDC 2019-nCoV EUA Kit method^26^. The RT-PCR was performed on the Applied Biosystems 7500 Fast Real-Time PCR Instrument with version 2.3 software.

In Study-2 and Study-3, two sets of randomly chosen ILC2 samples (see their preparation below) were analyzed using the procedure described above: the first set (Study-2) was analyzed prior to the shipment day and the second set (Study-3) was analyzed two days after the shipment day when the ILC2 participants started to test their samples.

Qualitative data (Supplemental Table 1A, 1C and 1D) indicated that all samples with RNA concentration at or above 50 copies per 50 μL were detected, and blank samples were not detected. The Ct values were subjected to quantitative analysis. There was no significant difference in the Ct values for the samples after 7, 14 and 21-day storage in Study 1 (Supplemental Table 1B), indicating the samples were stable for 21 days. The sample standard deviation (s_sample_) and standard deviation for the replicate measurements (s_e_) were in a range of 0-1.13 and 0.11-1.14, respectively, when they were calculated based on the Ct values. The homogeneity and stability results demonstrated that the trial samples were deemed sufficiently homogenous and stable, and the inoculation process was suitable to produce the targeted ILC2 samples.

### ILC2 sample preparation and pre-shipment tests

The RNA was inoculated into MTM in bulk, and 150 μL aliquots of each sample were transferred into 1.5-mL snap-top microfuge tubes, according to the sample composition in Table 1. All samples were stored at -80 ºC before shipping. To confirm successful inoculation before shipping, a set of ILC2 samples was tested as described in the Study-2 above. Another set of ILC2 samples was shipped to Cornell University to confirm the spiking levels by RT-ddPCR using the procedures described above.

**Table 1.**
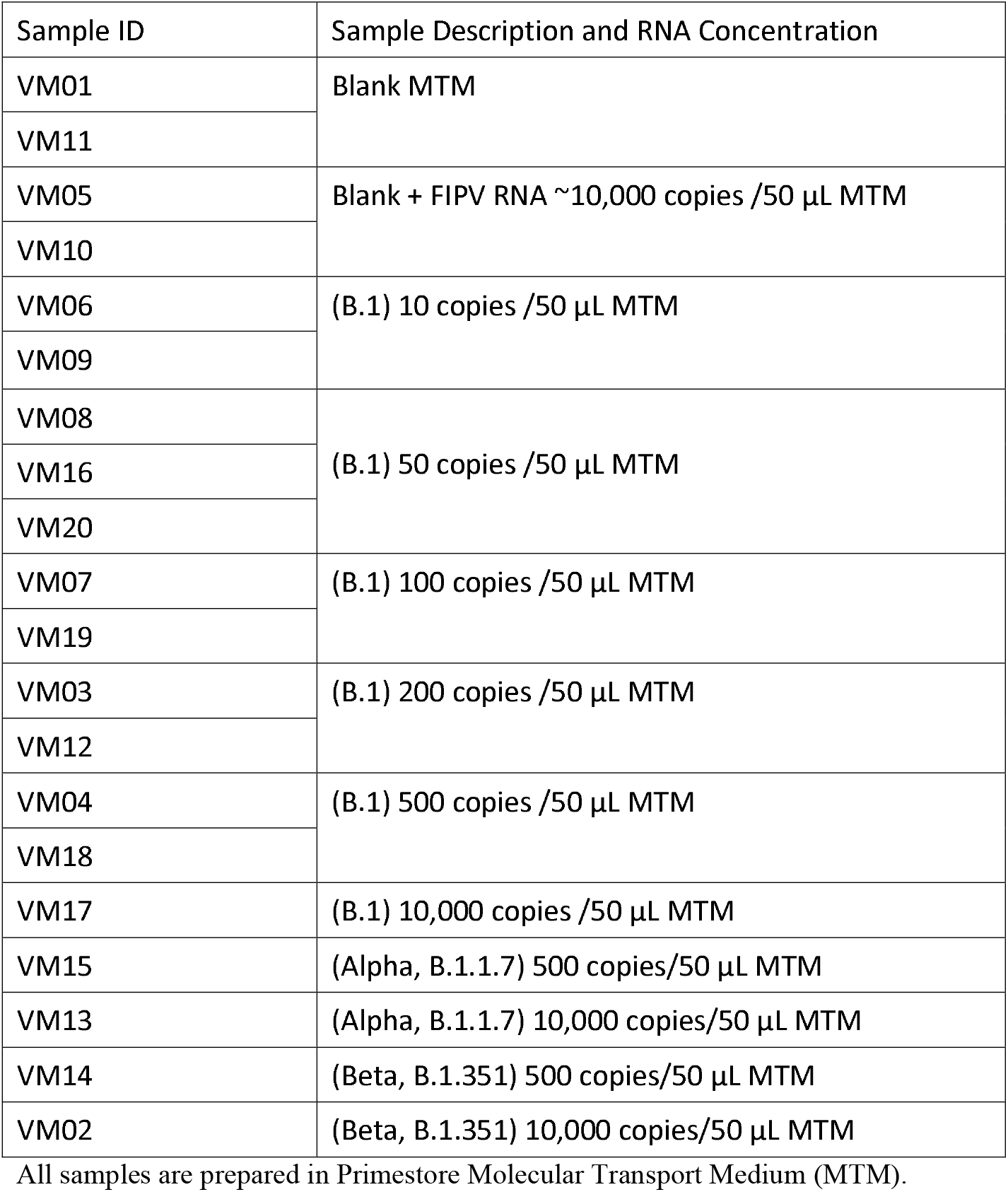
ILC2 samples description and RNA concentration.

| Sample ID | Sample Description and RNA Concentration |
| --- | --- |
| VM01 | Blank MTM |
| VM11 |  |
| VM05 | Blank + FIPV RNA ~10,000 copies /50 µL MTM |
| VM10 |  |
| VM06 | (B.1) 10 copies /50 µL MTM |
| VM09 |  |
| VM08 | (B.1) 50 copies /50 µL MTM |
| VM16 |  |
| VM20 |  |
| VM07 | (B.1) 100 copies /50 µL MTM |
| VM19 |  |
| VM03 | (B.1) 200 copies /50 µL MTM |
| VM12 |  |
| VM04 | (B.1) 500 copies /50 µL MTM |
| VM18 |  |
| VM17 | (B.1) 10,000 copies /50 µL MTM |
| VM15 | (Alpha, B.1.1.7) 500 copies/50 µL MTM |
| VM13 | (Alpha, B.1.1.7) 10,000 copies/50 µL MTM |
| VM14 | (Beta, B.1.351) 500 copies/50 µL MTM |
| VM02 | (Beta, B.1.351) 10,000 copies/50 µL MTM |

All samples are prepared in Primestore Molecular Transport Medium (MTM).

For the pre-shipment temperature trial, the packaging configuration was tested by packaging mock samples in dry ice in a shipping box according to the International Air Transport Association Dangerous Goods Regulations. After holding the box for 72 hours at room temperature, the sample containers were observed and qualitatively assessed as frozen or not. The assessment showed that the packaging configuration kept the primary sample containers frozen for 72 hours.

### ILC2 sample distribution

The final shipment samples were packaged using the Saf-T-Pak STP-320 UN 3373 Category B Frozen Insulated Shipping System according to the manufacturer-provided instructions and shipped via FedEx Priority Overnight. A total of 59 sets of blind-coded samples were shipped on dry ice to the 45 participating laboratories (14 laboratories requested a second set of samples to test two methods or to test by two analysts). Participants were not aware of spike levels (e.g., analyte concentration) or number of replicates per spike level prepared by the organizers.

### Sample analysis and data acquisition

Participants were instructed to use the SARS-CoV-2 RNA extraction and detection methods that they routinely use in their laboratories. To facilitate statistical analysis, all analysts used the same input volume (50 μl) for RNA extraction and reported their volumes of eluted RNA and PCR template to the ICL organizer. Sample handling and result reporting instructions were discussed with the participants via two training sessions. To ensure confidentiality, each analyst was assigned an analyst identification number (AIN). Each analyst reported the results as “detected” (D), “not detected” (N), or “inconclusive” (I) for SARS-CoV-2 viral RNA, according to their own laboratory’s protocols. Participants were also instructed to report Ct values for each PCR marker used, cut-off values, basic method information (e.g., PCR instrument model, extraction kit, and internal controls, as well as extraction, elution, and PCR input volumes), and any modifications to their methods. Detailed methods from each participating laboratory were kept confidential to maintain anonymity. We therefore refer to each assay target as “marker”, consistent with the terminology used in the ILC1 publication^20^.

### Qualitative and quantitative assessments

Rate of detection (ROD), the number of positive results divided by the total number of results, was calculated for all markers used (i.e., overall detection) and for N1 and N2 markers separately. Inconclusive results were classified as “not detected” for the assessment.

Analytical sensitivity (Level of Detection, LOD) was calculated based on a probability of detection (POD) model. The complementary log-log regression model^27,28^ (i.e., the statistical model that corresponds to the Poisson assumption) was modified to take into consideration the analyst-specific actual copy numbers per well for calculating the marker- and analyst-specific POD curves. The POD curve was calculated based on ROD values obtained from the original (i.e., not rounded) Ct values. From these POD curves, the LOD95 values (the numbers of copies at which a POD of 95% is achieved) were derived. LOD95 was based on the effective volume (i.e., that part of the original sample volume used in the RT-PCR), which was calculated based on three volumes (e.g., extraction, elution, and PCR input volumes) reported by participants to organizers (hereafter referred to as LOD95_eff. vol_). The adjustment was necessary for meaningful evaluation of method sensitivities for the individual laboratories.

The PCR amplification rate (i.e., efficiency) was calculated separately for each marker based on the nominal copy numbers (equivalent to dilution levels) and the submitted Ct values.

## Results

### Overall detection results

Fifty-seven (57) datasets submitted from 45 laboratories were collected and analyzed. The analysts submitted qualitative “overall detection” results (Table 2), Ct values for various markers (Supplemental Tables 2-6), Ct cut-off values (Table 3), and basic method information. The analysts targeted different markers for the SARS-CoV-2 RNA testing. The “overall detection” results and Ct cut-off values reported are based on criteria selected by individual analysts according to their internal protocols. Comparative evaluation of qualitative “overall detection” results, reported Ct values and the Ct cut-off values revealed that individual analysts used different decision-making criteria during interpretation of their datasets (Table 2). Specifically, some analysts reported Detected (D), whereas other analysts reported Not Detected (N) or Inconclusive (I) when some, but not all, targeted markers were detected (i.e., one marker was detected out of two targeted markers or three markers were detected out of four targeted markers).

**Table 2.**
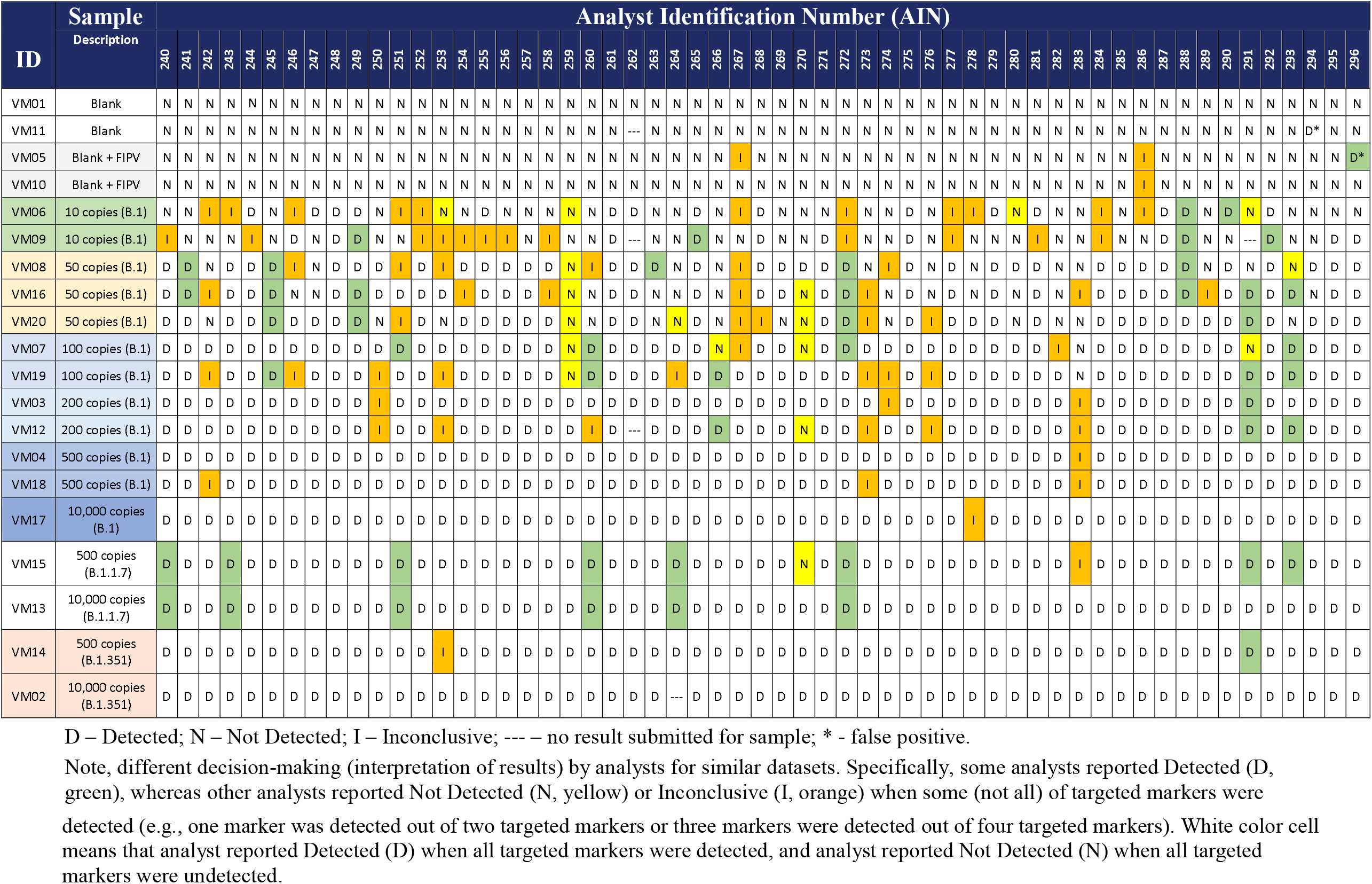
The “overall detection” results submitted by all participants for ILC2 samples.

**Table 3.** Sensitivity (LOD95_eff. Vol_.) and PCR efficiency for each marker

| AIN | Effective volume<sup>†</sup><br>(μl) | Ct Cut Off Values | LOD95<sub>eff. volume</sub><sup>‡</sup> and efficiency (%) |  |  |  |  |  |  |  |
| --- | --- | --- | --- | --- | --- | --- | --- | --- | --- | --- |
|  |  |  | N1 | N2 | N | N3 | E | RdRP | ORF1ab | S |
| 278 | 0.5 | 45 | 2.0 ** (210%) | 1.9 ** (106%) |  |  |  |  |  |  |
| 276 | 0.5 | 40 | 3.1 (117%) | 4.6 (115%) |  |  |  |  |  |  |
| 263 | 1.1 | 40 | 12.7 (114%) | 4.4 (106%) |  |  |  |  |  |  |
| 293 | 2.5 | 40 | 14.1 (260%) | 6.4 (158%) |  |  |  |  |  |  |
| 291 | 2.5 | 40 | 16.8 (193%) | 14.9 (155%) |  |  |  |  |  |  |
| 285 | 2.5 | 45 | 3.3 (107%) | 3.3 (83%) |  |  |  |  |  |  |
| 247 | 2.5 | 45 |  |  |  |  |  | 7.4 (140%) |  |  |
| 250 | 2.5 | 40 |  |  | 26.7 (204%) |  |  |  |  |  |
| 262 | 2.5 | 40 |  |  | 6.6 * (99%) |  |  |  |  |  |
| 294 | 2.5 | 40 |  |  | 4.9 (85%) |  |  |  |  |  |
| 283 | 2.8 | 36 | >556 (87%) | 106.7 (101%) |  |  |  |  |  |  |
| 289 | 2.8 | 40 | 10.9 (106%) | 7.1 (104%) |  |  |  |  |  |  |
| 241 | 2.8 | 39.99 | 10.9 (88%) | 3.7 (102%) |  |  |  |  |  | 18.2 (92%) |
| 280 | 2.8 | 40 | 10.9 (97%) | 8.2 (90%) |  |  |  |  |  |  |
| 246 | 2.8 | 40 | 13.2 (111%) | 3.7 (120%) |  |  |  |  |  |  |
| 282 | 2.8 | 40 | 15.7 (101%) | 22.9 (79%) |  |  |  |  |  |  |
| 267 | 2.8 | 40 | 17 (125%) | 6.4 (201%) |  |  |  |  |  |  |
| 279 | 2.8 | 40 | 2.0 ** (122%) | 2.0 ** (108%) |  |  |  |  |  |  |
| 265 | 2.8 | 40 | 2.0 ** (94%) | 4.6 * (80%) |  |  |  |  |  |  |
| 258 | 2.8 | 40 | 2.0 ** (96%) | 7.1 (93%) |  |  |  |  |  |  |
| 245 | 2.8 | 45 | 22.9 (112%) | 3.7 (103%) |  |  |  |  |  |  |
| 257 | 2.8 | 45 | 3.7 (106%) | 3.7 (92%) |  |  |  |  |  |  |
| 256 | 2.8 | 40 | 3.7 (91%) | 2.0 ** (100%) |  |  |  |  |  |  |
| 249 | 2.8 | 45 | 3.7 (92%) |  | 5.2 (132%) |  |  |  |  |  |
| 274 | 2.8 | 45 | 34.1 (154%) | 10.9 (129%) |  |  |  |  |  |  |
| 261 | 2.8 | 40 | 4.6 * (101%) | 4.6 * (111%) |  |  |  |  |  |  |
| 284 | 2.8 | 40 | 4.6 * (101%) | 4.6 * (86%) |  |  |  |  |  |  |
| 255 | 2.8 | 45 | 4.6 * (95%) | 2.0 ** (67%) |  | 2.0 ** (99%) |  |  |  |  |
| 242 | 2.8 | 40 | 41.8 (114%) | 8.2 (100%) |  |  |  |  |  |  |
| 273 | 2.8 | 40 | 55.9 (107%) | 7.1 (108%) |  |  |  |  |  |  |
| 252 | 2.8 | 40 |  |  |  |  | 3.7 (88%) | 4.6 * (111%) |  |  |
| 269 | 3.1 | 40 | 8 (112%) | 8 (134%) |  |  |  |  |  |  |
| 275 | 3.9 | 45 |  |  | 6.5 * (106%) |  |  |  |  |  |
| 271 | 4.2 | 40 | 3 (104%) | 3 (91%) |  |  |  |  |  |  |
| 244 | 4.2 | 40 | 3 (110%) | 3 (99%) |  |  |  |  |  |  |
| 266 | 4.4 | 37 | 36.6 (136%) | 104 (84%) |  |  |  |  |  |  |
| 254 | 5 | 40 | 12.8 (119%) | 9.3 (109%) |  |  |  |  |  |  |
| 290 | 5 | 40 | 6.7 (102%) | 3.6 (108%) |  |  |  |  |  |  |
| 268 | 5 | 45 | 6.7 (110%) | 8.3 * (108%) |  |  |  |  |  |  |
| 253 | 5 | 45 | 66.8 (72%) | 9.3 (107%) |  |  |  |  |  |  |
| 259 | 5 | 40 |  |  |  |  | 3.6 (135%) | 57 (100%) |  |  |
| 270 | 5 | 40 |  |  |  |  | 6.7 (102%) | 16 (93%) |  |  |
| 286 | 5 | 45 |  |  |  |  | 6.7 (126%) |  | 2.4 ** (84%) |  |
| 248 | 5.6 | 45 |  |  | 9.3 * (138%) |  |  |  |  |  |
| 287 | 5.8 | 38 |  |  | 9.7 * (108%) |  |  |  |  |  |
| 240 | 6.7 | 37 |  |  | 29.6 (94%) |  |  |  | 4.8 (115%) | 52.9 (88%) |
| 295 | 7 | 40 |  |  | 3.2 (92%) |  |  |  |  |  |
| 288 | 8 | 36 |  |  |  |  | 13.3 * (103%) | 31.4 (101%) |  |  |
| 292 | 8 | 36 |  |  |  |  | 13.3 * (126%) | 5.7 (101%) |  |  |
| 277 | 8.3 | 40 | 6 (117%) | 6 (99%) |  |  |  |  |  |  |
| 296 | 8.9 | 40 | 6.4 (104%) | 3.7 (111%) |  |  |  |  |  |  |
| 243 | 10 | 37 |  |  | 25.6 (107%) |  |  |  | 7.2 (121%) | 79.3 (85%) |
| 251 | 10 | 37 |  |  | 56.4 (119%) |  |  |  | 7.2 (115%) | >2000 (65%) |
| 260 | 10 | 37 |  |  | 91.6 (73%) |  |  |  | 122.7 (64%) | >2000 (59%) |
| 264 | 10 | 37 |  |  | 82.5 (88%) |  |  |  | 56.4 (94%) | >2000 (70%) |
| 272 | 10 | 39 |  |  | 44.4 (97%) |  |  |  | 13.3 (108%) | >2000 (93%) |
| 281 | 11.1 | 40 | 18.5 * (110%) | 8 (104%) |  |  |  |  |  |  |

<sup>†</sup> Effective volume is that part of the original sample volume used in the PCR by each analyst. It was calculated based on three volumes (e.g., extraction, elution and PCR input) used and reported by analysts.
<sup>‡</sup> LOD<sub>95 eff. vol.</sub> is the number of copies in a PCR reaction at which a POD of 95% is achieved based on the effective volume used.
\* Upper limit of confidence interval as there are no false negative results.
\*\* Value is not significantly lower than the theoretical optimum of 3 target copies.

The ROD was calculated and summarized at each spike level (Table 4). For the overall detection as well as for the two most common markers (N1 and N2), the ROD values increased with increasing copy numbers as expected, consistently achieving ROD values above 95% at 500 copies / 50 μL. At 100 copies / 50 μL, overall detection was 85%. Lower copy levels were also included to help assess levels of detection, and at the lowest spike level of 10 copies/ 50 μL, 26% of samples were still identified as positive for SARS-CoV-2. One exception was observed where for B.1 samples at 10,000 copies per 50 μL, the ROD for N2 marker was less than 100% (i.e., 97%). This was due to one false negative result submitted at this high level, which also affected the calculated value of LOD95_eff. vol._ for the analyst (AIN 278).

**Table 4.** Rate of detection (ROD) calculated for all participants (e.g., overall detection) and for those who used N1 and N2 markers

| SARS-CoV-2 RNA spiking level | Number of PCR replicates | Rate of detection (ROD) across analysts* |  |  |
| --- | --- | --- | --- | --- |
|  |  | Overall detection | N1 | N2 |
| Number of analysts included in the calculations |  | 57 | 37 | 36 |
| Blank | 2 | 1% | 0% | 0% |
| Blank + 10,000 FIPV copies / 50 µL | 2 | 1% | 0% | 3% |
| B.1 10 copies / 50 µL | 2 | 26% | 30% | 35% |
| B.1 50 / 50 µL | 3 | 65% | 56% | 70% |
| B.1 100 copies / 50 µL | 2 | 85% | 81% | 92% |
| B.1 200 copies / 50 µL | 2 | 91% | 95% | 96% |
| B.1 500 copies / 50 µL | 2 | 96% | 95% | 100% |
| B.1 10,000 copies / 50 µL | 1 | 98% | 100% | 97% |
| Alpha (B.1.1.7) 500 copies / 50 µL | 1 | 96% | 97% | 100% |
| Alpha (B.1.1.7) 10,000 copies / 50 µL | 1 | 100% | 100% | 100% |
| Beta (B.1.351) 500 copies / 50 µL | 1 | 98% | 97% | 100% |
| Beta (B.1.351) 10,000 copies / 50 µL | 1 | 100% | 100% | 100% |

\* ROD values were calculated based on the original (i.e., not rounded) Ct values. Inconclusive results and results with Ct values higher than the analyst-specific cut-off were classified as “not detected” in this statistical summary.

We also sought to determine whether ROD varied for different SARS-CoV-2 variants. For the Alpha (B.1.1.7) and Beta (B.1.351) variants, ROD values for N1, N2, and overall detection were nearly identical to the results from B.1, indicating laboratories were able to detect SARS-CoV-2 regardless of the variant. Among blank samples, there was only one false positive (AIN 294), indicating a false positive rate of < 1% for all participants.

### Analytical Specificity of methods

The analytical specificity of the methods was evaluated by including the feline infectious peritonitis virus (FIPV) coronavirus RNA as a confounder. As shown in Table 2, only one analyst (AIN 296) reported one false positive for one of the two “Blank + FIPV” samples. Importantly, this analyst detected the N2 marker, but not the N1 marker for this sample (see Supplemental Table 2) and thus reported the sample as “Detected” according to their internal protocols. Similarly, two other analysts (AIN 267 and 286) detected the “Blank + FIPV” samples with only one of the markers (see Supplemental Table 2 and 5), however, they reported the sample as “Inconclusive” according to their internal protocols. Overall, ILC2 results revealed that participants’ methods are specific to SARS-CoV-2 and do not routinely yield false positive results for FIPV.

### Analytical sensitivity and efficiency of methods

Analytical sensitivity was evaluated with LOD95_eff. vol._ (Table 3), which was calculated based on the actual RNA copy number added to the PCR for each individual analyst (i.e., effective volume for each spiking level). The effective volumes tested varied by a factor of 22 among analysts and ranged from 0.5 to 11.1 μL (Table 3). The LOD95_eff. vol._ values greatly varied among participants and markers they used. Sensitivities for N1 and N2 markers were summarized (Table 5), including LOD95_eff. vol._ values calculated for AIN 283 and AIN 266, which may be considered as outliers. Specifically, AIN 283 and AIN 266 reported Ct values for multiple samples at low spike levels (≤ 200 copies) (Supplemental Table 2). However, those values were counted as not detected due to being higher than the Ct cut-off values established by the analysts. This indicates that the methods in these two laboratories are likely sensitive enough to detect more samples at lower spike levels, but established Ct cut-off values were too stringent (i.e., unoptimized) resulting in more false negative results (see Table 2) affecting LODs in these two laboratories. Thus, LOD95_eff. vol._ calculations with these two laboratories (AIN 283 and 266 for N1 and N2) are shown in Table 5; however, LOD95_eff. vol._ with these two laboratories excluded are interpreted below.

**Table 5.** Comparative summary of sensitivity per each marker

| Marker | Total number of analysts | No. of analysts without false negative results | Median LOD<sub>95<sub>eff. vol.</sub></sub> | Minimum LOD<sub>95<sub>eff. vol.</sub></sub> | Maximum LOD<sub>95<sub>eff. vol.</sub></sub> | Number of analysts with LOD<sub>95<sub>eff. vol.</sub></sub> at ≤ 20 copies |
| --- | --- | --- | --- | --- | --- | --- |
| N1 | 37 | 4 | 8.0 | 2.0 | >556 | 30/37 (81%) |
| N1* | 36 | 4 | 6.7 | 2.0 | 66.8 | 30/36 (83%) |
| N2 | 36 | 4 | 5.3 | 1.9 | 106.7 | 33/36 (92%) |
| N2** | 34 | 4 | 4.6 | 1.9 | 22.9 | 33/34 (97%) |
| E | 6 | 2 | 6.7 | 3.6 | 13.3 | 6/6 (100%) |
| N | 14 | 4 | 17.7 | 3.2 | 91.6 | 7/14 (50%) |
| N3 | 1 | --- | --- | --- | --- | 1/1 (100%) |
| ORF1ab | 7 | 0 | 7.2 | 2.4 | 122.7 | 5/7 (71%) |
| RdRP | 6 | 1 | 11.7 | 4.6 | 57.0 | 4/6 (67%) |
| S | 7 | 0 | 52.9 | 18.2 | 79.3 | 1/7 (14%) |

\* An LOD<sub>eff. vol.</sub> value of >556 copies by AIN 283 was excluded from the summary because this value was considered as an outlier due to unoptimized (i.e., too stringent) Ct cut-off values used by the analyst and substantially affecting the summary of LODs for N1 marker.
\*\* LOD<sub>95<sub>eff. vol.</sub></sub> values of 106 copies (AIN 283) and 104 copies (AIN 266) were excluded from calculations for the same reason described above.

As mentioned above, LOD95_eff. vol._ values from participants were combined for each marker and their median, minimum, and maximum LOD95_eff. vol._ values for comparative evaluation (Table 5). The LOD95_eff. vol._ values ranged from 2.0 to 66.8 for the N1 marker, with a median of 6.7. This corresponded to a factor of around 30 between the lowest and highest values; while some analysts detected every copy of SARS-CoV-2 RNA, others only detected less than 10% of the copies. For the N2 marker, the LOD95_eff. vol._ values ranged from 1.9 to 22.9 with the median value of 4.6 copies indicating that methods based on N2 marker were the most sensitive in ILC2. An ideal LOD95_eff. vol._ is calculated to be 3 copies per PCR reaction based on a hypothetical POD curve^28^. Due to random variation, LOD95_eff. vol._ values below 3 may be observed. For the N1 marker, 30 out of 36 analysts (83%) had an LOD95_eff. vol._ value ≤20 copies and 20 out of 37 analysts (54%) had an LOD95_eff. vol._ value not statistically significantly greater than 3 (meaning that the LOD95_eff. vol._ is within the margin of error of the best possible value). For the N2 marker, 33 out of 34 analysts (97%) had an LOD95_eff. vol._ value ≤20 copies and 27 out of 36 analysts (75%) had an LOD95_eff. vol._ value not statistically significantly greater than 3. For the E and N3 markers, all analysts (100%) had LOD95_eff. vol._ values at ≤20 copies. For the N, ORF1ab and RdRP markers, only half of the analysts had LOD95_eff. vol._ near or equal to the best possible LOD – theoretical minimum of 3 copies. For the S marker, none of the analysts had LOD95_eff. vol._ near or equal to the best possible value of 3 copies and only 1 out of 7 analysts (14%) had an LOD95_eff. vol._ below 20 copies.

Calculated efficiency greatly varied among participants and markers they used as well (Table 3). In general, PCR efficiency of 100% indicates that the target sequence of interest doubles during each cycle. If the Ct values change less than 3.3 cycles between 10-fold dilutions of the PCR template, it resulted in efficiency values that were greater than 100%. Some of the calculated efficiencies are indeed higher than 100%, which could be interpreted as an indication of problems in the amplification process. On the other hand, it should be noted here that the calculation of efficiencies is associated with considerable statistical uncertainty due to unavoidable random fluctuations in the Ct values. This is especially true when - as in the present case - the underlying dilution levels differ by only a few orders of magnitude.

## Discussion

The ILC2 provides insight into performance of methods to detect SARS-CoV-2 RNA in PrimeStore molecular transport medium. ILC2 demonstrated that most participants have relatively sensitive and specific methods to detect three SARS-CoV-2 variants.

The testing methods varied between laboratories, as the participating analysts were instructed to follow their routine SARS-CoV-2 detection procedure for ILC2. This, plus the variability in the laboratory’s Ct cut-off values, resulted in some inconsistency in interpretations among different analysts. For a study such as ILC2 where various extraction and detection methods were involved, applying a universal Ct cut-off value for each marker to provide a score for each individual analyst is not realistic. Method information provided by each analyst allowed the ILC-2 organizers to statistically identify possible correlations with result variability. Analyst-specific results broken down by Ct values for different markers and with extraction and PCR methods annotated were summarized and provided to the analysts in an ILC2 report. These data are not shown in this manuscript to protect confidentiality of participants, particularly those who were using methods unique to their laboratory.

Most of the participating laboratories used the CDC N1 and N2 assays for detecting the SARS-CoV-2 RNA. Research has reported one marker as more sensitive than the other or vice versa^29,30^. In the ILC1^20^, differences in ROD values between the two markers were very minor. In the current ILC2, the ROD values were higher for N2 than for N1, especially for 50 to 100 copies/50 μL for the B.1 variant. The LOD95_eff. vol._ values generated for the N1 and N2 markers indicated that 81% and 92% of ILC2 analysts, respectively, demonstrated an LOD95_eff. vol._ value below 20 copies.

The relationship between the effective volumes used and the observed LOD95_eff. vol._ values was not significant (Table 3). While it cannot be ruled out that high effective volumes make it more difficult to detect all copies, the observed LOD95_eff. vol._ values also depend on other factors. These factors may include extraction kit, PCR kit, reagents, model/type of equipment such as PCR machine, centrifuges and pipettes, level of analyst’s experience, quality control system in laboratory, multiplex versus single-plex approaches and others. Information on some of these factors was provided in the confidential report for the participants. The variability in Ct values observed for particular analysts is also informative and can point to potential issues with methods that may affect success of analysts in future exercises (Supplemental Tables 2-6).

The ILC2 study demonstrated a successful collaboration involving government agencies, universities, and private industry. By using a larger range of SARS-CoV-2 RNA spiking levels, including lower concentrations, the LOD95_eff. vol._ values of the methods used by participating analysts were evaluated more accurately than we did in ILC1. The laboratories were able to detect Alpha and Beta variants of SARS-COV-2 with their current methods. The method specificity was confirmed by using FIPV RNA as a confounder and reached over 99%. In summary, the ILC2 was a success in meeting the stated objectives. The exercise allowed organizers not only to characterize important parameters of participants’ method performance (e.g., analytical sensitivity and specificity, efficiency, and suitability for different variants) but also allowed participants to compare their performance to each other. Organizers processed the submitted data using various statistical approaches that allowed them to identify possible weaknesses and strengths of methods used, and offer suggestions on improving participants’ performance in the future. Specifically, an important finding of the ILC2 is that individual analysts used different decision-making criteria during interpretation of similar datasets. This indicates a need for laboratories to review data from this exercise and potentially reassess their decision-making criteria during interpretation of Ct values when using multiple markers. The ILC2 study also indicates that the false negative rate and sensitivity of some methods can be improved if Ct cut-off values used are re-evaluated (e.g., on a test that a too stringent Ct cut-off value was originally used) and optimized by analysts accordingly. In the current era of rapidly developing methodology and lack of international standards, participation in ILCs like this study is very beneficial. In contrast to other types of proficiency testing exercises that only aim to assess which results are correct or incorrect, this ILC revealed much more about the methods used and assist participants in continuous efforts to improve performance.

## Data Availability

All data produced in the present work are contained in the manuscript

## Acknowledgments

We acknowledge the diligence and hard work of the laboratory scientists who rapidly developed SARS-CoV-2 assays for animals and participated in this collaborative study.

We acknowledge the following individuals for technical assistance and administrative support: Angelica Jones, Olgica Ceric, Ellen Hart, and Dave Rotstein from FDA, Joseph Flint from Cornell University, and Mia Torchetti and Christina Loiacono from USDA. We also thank Kirsten Simon from QuoData for supporting and facilitating this work. The Genomics Facility of the Biotechnology Resource Center at the Cornell University Institute of Biotechnology assisted with copy number quantification, and we thank Peter Schweitzer for facilitating this. We also thank Robert Newkirk from FDA CFSAN Proficiency Team for sample shipment assistance.

## Disclaimer

The views expressed in this article are those of the authors and do not necessarily reflect the official policy of the Department of Health and Human Services, the U.S. Food and Drug Administration, or the U.S. Government. The use of trade, firm, or product names is for descriptive purposes only and does not imply endorsement by the U.S. Government.

## Supplemental material

Supplemental material for this article is available online.

## References

1. Ksiazek TG, Erdman D, Goldsmith CS, et al. A novel coronavirus associated with severe acute respiratory syndrome. N Engl J Med 2003;348:1953–1966.

2. Coleman CM, Frieman MB. Emergence of the Middle East respiratory syndrome coronavirus. PLoS Pathog 2013;9:e1003595.

3. Oude Munnink BB, Sikkema RS, Nieuwenhuijse DF, et al. Jumping back and forth: anthropozoonotic and zoonotic transmission of SARS-CoV-2 on mink farms. bioRxiv 2020.

4. Oude Munnink BB, Sikkema RS, Nieuwenhuijse DF, et al. Transmission of SARS-CoV-2 on mink farms between humans and mink and back to humans. Science 2021;371:172–177.

5. He S, Han J, Lichtfouse E. Backward transmission of COVID-19 from humans to animals may propagate reinfections and induce vaccine failure. Environ Chem Lett 2021;3:1–6.

6. Kumar R, Harilal S, Al-Sehemi AG, et al. COVID-19 and domestic animals: exploring the species barrier crossing, zoonotic and reverse zoonotic transmission of SARS-CoV-2. Curr Pharm Des 2021;27:1194–1201.

7. Anonymous. Animals and COVID-19. In. https://www.cdc.gov/coronavirus/2019-ncov/daily-life-coping/animals.html: Centers for Disease Control and Prevention; 2021.

8. Newman A, Smith D, Ghai RR, et al. First reported cases of SARS-CoV-2 infection in companion animals - New York, March-April 2020. MMWR Morb Mortal Wkly Rep 2020;69:710–713.

9. Hamer SA, Pauvolid-Corrêa A, Zecca IB, et al. Natural SARS-CoV-2 infections, including virus isolation, among serially tested cats and dogs in households with confirmed human COVID-19 cases in Texas, USA. bioRxiv 2020.

10. McAloose D, Laverack M, Wang L, et al. From people to panthera: natural SARS-CoV-2 infection in tigers and lions at the Bronx zoo. mBio 2020;11.

11. Lee AC, Zhang AJ, Chan JF, et al. Oral SARS-CoV-2 inoculation establishes subclinical respiratory infection with virus shedding in golden Syrian hamsters. Cell Rep Med 2020;1:100121.

12. Meisner J, Baszler TV, Kuehl KH, et al. Household transmission of SARS-CoV-2 from humans to dogs in Washington and Idaho: burden and risk factors. bioRxiv 2021.

13. Ruiz-Arrondo I, Portillo A, Palomar AM, et al. Detection of SARS-CoV-2 in pets living with COVID-19 owners diagnosed during the COVID-19 lockdown in Spain: A case of an asymptomatic cat with SARS-CoV-2 in Europe. Transbound Emerg Dis 2021;68:973–976.

14. Sailleau C, Dumarest M, Vanhomwegen J, et al. First detection and genome sequencing of SARS-CoV-2 in an infected cat in France. Transbound Emerg Dis 2020;67:2324–2328.

15. Segales J, Puig M, Rodon J, et al. Detection of SARS-CoV-2 in a cat owned by a COVID-19-affected patient in Spain. Proc Natl Acad Sci U S A 2020;117:24790–24793.

16. U.S. Department of Agriculture. Cases of SARS-CoV-2 in Animals in the United States,. In. https://www.aphis.usda.gov/animal_health/one_health/downloads/sars-cov2-in-animals.pdf.: 2020.

17. Wacharapluesadee S, Tan CW, Maneeorn P, et al. Evidence for SARS-CoV-2 related coronaviruses circulating in bats and pangolins in Southeast Asia. Nat Commun 2021;12:972.

18. Ip HS, Griffin KM, Messer JD, et al. An opportunistic survey reveals an unexpected coronavirus diversity hotspot in North America. Viruses 2021;13:2016–2028.

19. Kaneene JB, Warnick LD, Bolin CA, et al. Changes in multidrug resistance of enteric bacteria following an intervention to reduce antimicrobial resistance in dairy calves. J Clin Microbiol 2009;47:4109–4112.

20. Deng K, Uhlig S, Ip HS, et al. Interlaboratory comparison of SARS-CoV2 molecular detection assays in use by U.S. veterinary diagnostic laboratories. J Vet Diagn Invest 2021.

21. Badua C, Baldo KAT, Medina PMB. Genomic and proteomic mutation landscapes of SARS-CoV-2. J Med Virol 2021;93:1702–1721.

22. Galloway SE. Emergence of SARS-CoV-2 B.1.1.7 Lineage — United States, December 29, 2020–January 12, 2021. MMWR 2021;70:95–99.

23. Tegally H, Wilkinson E, Giovanetti M, et al. Detection of a SARS-CoV-2 variant of concern in South Africa. Nature 2021;592:438–443.

24. O’Brien A, Mettelman RC, Volk A, et al. Characterizing replication kinetics and plaque production of type I feline infectious peritonitis virus in three feline cell lines. Virology 2018;525:1–9.

25. Dye C, Helps CR, Siddell SG. Evaluation of real-time RT-PCR for the quantification of FCoV shedding in the faeces of domestic cats. J Feline Med Surg 2008;10:167–174.

26. Anonymous. CDC 2019-novel coronavirus (2019-nCoV) real-time RT-PCR diagnostic panel for emergency use only instructions for use. In. https://www.fda.gov/media/134922/download: Centers for Disease Control and Prevention; 2020.

27. International Organization for Standardization. Microbiology of the food chain -- Method validation -- Part 2: Protocol for the validation of alternative (proprietary) methods against a reference method. In: ISO 16140-2:20162016.

28. Uhlig S, Frost K, Colson B, et al. Validation of qualitative PCR methods on the basis of mathematical statistical modelling of the probability of detection. Accreditation and Quality Assurance 2015;20:75–83.

29. Vogels CBF, Brito AF, Wyllie AL, et al. Analytical sensitivity and efficiency comparisons of SARS-CoV-2 RT-qPCR primer-probe sets. Nat Microbiol 2020;5:1299–1305.

30. Perchetti GA, Nalla AK, Huang ML, et al. Validation of SARS-CoV-2 detection across multiple specimen types. J Clin Virol 2020;128:104438.

