## Supplemental tables for "Second round of the interlaboratory comparison (ILC) exercise of SARS-CoV-2 molecular detection assays being used by 45 veterinary diagnostic laboratories in the US"

Any use of trade, firm, or product names is for descriptive purposes only and does not imply endorsement by the U.S. Government.

**Supplemental Table 1. Stability and homogeneity results from Study-1, 2, and 3.**

**(A) Results from Study-1: Trials for stability and homogeneity of RNA in PrimeStore Molecular Transport Medium (MTM)**

For each analyst, three sets (A-C) of 18 samples described above at levels of 0, 10, 20, 50, 100, or 1,000 copies/50 µL MTM were tested on Day 0, 7, and 14 of storage at -80 °C. A set of 6 samples at level of 100 to 1,000 copies/50 µL MTM was tested on Day 21 by Analyst 2. The trial samples were inoculated and stored in the same manner as shipment samples.

| **Sample set** | **Sample** | **SARS-CoV-2 RNA Inoculation Level (copies/50 µL)** | **Analyst 1 Results** | **Analyst 2 Results** |
|---|---|---|---|---|
| **A** | Day 0 – S1 | 0 | ND | ND |
| | Day 0 – S2 | 10 | D | D |
| | Day 0 – S3 | | D | D |
| | Day 0 – S4 | | D | D |
| | Day0 – S5 | | D | D |
| | Day 0 – S6 | 20 | D | D |
| | Day 0 – S7 | | D | D |
| | Day 0 – S8 | | D | D |
| | Day 0 – S9 | | D | D |

| | | | | |
|---|---|---|---|---|
| | Day 0 – S10 | 50 | D | D |
| | Day 0 – S11 | | D | D |
| | Day 0 – S12 | | D | D |
| | Day 0 – S13 | 100 | D | D |
| | Day 0 – S14 | | D | D |
| | Day0 – S15 | 1000 | D | D |
| | Day 0 – S16 | | D | D |
| | Day 0 – S17 | Alpha (B.1.1.7)- 1000 | D | D |
| | Day 0 – S18 | Beta (B.1.351)- 1000 | D | D |
| **B** | Day 7 – S1 | 0 | ND | ND |
| | Day 7 – S2 | 10 | D | D |
| | Day 7 – S3 | | D | D |
| | Day 7 – S4 | | D | D |
| | Day 7 – S5 | | D | D |
| | Day 7 – S6 | 20 | D | D |
| | Day 7 – S7 | | D | D |
| | Day 7 – S8 | | ND | D |
| | Day 7 – S9 | | D | D |
| | Day 7 – S10 | 50 | D | D |
| | Day 7 – S11 | | D | D |
| | Day 7 – S12 | | D | D |
| | Day 7 – S13 | 100 | D | D |
| | Day 7 – S14 | | D | D |
| | Day 7 – S15 | 1000 | D | D |

| | | | | |
|---|---|---|---|---|
| | Day 7 – S16 | | D | D |
| | Day 7 – S17 | Alpha (B.1.1.7)- 1000 | D | D |
| | Day 7– S18 | Beta (B.1.351)- 1000 | D | D |
| **C** | Day 14 – S1 | 0 | ND | ND |
| | Day 14 – S2 | 10 | D | D |
| | Day 14 – S3 | | D | D |
| | Day 14 – S4 | | D | D |
| | Day 14 – S5 | | D | D |
| | Day 14 – S6 | 20 | D | D |
| | Day 14 – S7 | | D | D |
| | Day 14 – S8 | | D | D |
| | Day 14 – S9 | | D | D |
| | Day 14 – S10 | 50 | D | D |
| | Day 14 – S11 | | D | D |
| | Day 14 – S12 | | D | D |
| | Day 14 – S13 | 100 | D | D |
| | Day 14 – S14 | | D | D |
| | Day 14 – S15 | 1000 | D | D |
| | Day 14 – S16 | | D | D |
| | Day 14 – S17 | Alpha (B.1.1.7)- 1000 | D | D |
| | Day 14 – S18 | Beta (B.1.351)- 1000 | D | D |
| **D** | Day 21 – S13 | 100 | N/A | D |
| | Day 21 – S14 | | N/A | D |
| | Day 21 – S15 | 1000 | N/A | D |

| | | | | |
|---|---|---|---|---|
| | Day 21 – S16 | | N/A | D |
| | Day 21 – S17 | Alpha (B.1.1.7)- 1000 | N/A | D |
| | Day 21 – S18 | Beta (B.1.351)- 1000 | N/A | D |

Note: Each set of Study-1 samples was tested by using Qiagen RNeasy Mini kit for manual RNA extraction. The RNA was isolated from the 50 µL samples and 30 µL was eluted from the Qiagen RNeasy mini kit purification column. The purified RNA (5 µL) was analyzed using AgPath-ID One-Step RT-PCR Kit.

S: Sample. D: Determined by either N1 and/or N2 marker; ND: not determined by both N1 and N2 markers; N/A: not available.

### (B) Statistical result of stability test

| **Analyst** | **Marker** | **Test Day** | **Number of data points** | **Regression Parameters*** | | **Efficiency [%]** | **Expected Ct Values Number of Copies per Reaction** | | | | |
|---|---|---|---|---|---|---|---|---|---|---|---|
| | | | | **Intercept** | **Slope** | | **10** | **100** | **1000** | **10000** | **LOD95**** |
| **1** | **N1** | 0 | 8 | 37.31 | -2.50 | 152 | 34.81 | 32.32 | 29.82 | 27.32 | 15 |
| | | 7 | 18 | 37.89 | -3.08 | 111 | 34.81 | 31.72 | 28.64 | 25.56 | 10 |
| | | 14 | 19 | 38.24 | -3.10 | 110 | 35.14 | 32.04 | 28.93 | 25.83 | 9 |
| | | 21 | 18 | 37.55 | -2.85 | 124 | 34.70 | 31.84 | 28.99 | 26.13 | 9 |
| | **N2** | 0 | 6 | 40.22 | -3.64 | 88 | 36.58 | 32.94 | 29.31 | 25.67 | >25 |
| | | 7 | 20 | 40.29 | -3.65 | 88 | 36.64 | 32.99 | 29.34 | 25.69 | 5 |
| | | 14 | 20 | 39.71 | -3.76 | 85 | 35.95 | 32.19 | 28.44 | 24.68 | 5 |
| | | 21 | 19 | 39.74 | -3.72 | 86 | 36.02 | 32.30 | 28.57 | 24.85 | 6 |
| **2** | **N1** | 0 | 11 | 36.53 | -2.13 | 194 | 34.40 | 32.26 | 30.13 | 28.00 | 5 |
| | | 7 | 18 | 38.19 | -2.86 | 124 | 35.33 | 32.47 | 29.61 | 26.76 | 9 |
| | | 14 | 21 | 38.31 | -3.08 | 111 | 35.23 | 32.16 | 29.08 | 26.00 | 5 |

| | | | | | | | | | | | |
|---|---|---|---|---|---|---|---|---|---|---|---|
| | | 21 | 14 | 37.06 | -2.08 | 202 | 34.98 | 32.90 | 30.82 | 28.74 | 17 |
| | **N2** | 0 | 8 | 38.85 | -2.59 | 143 | 36.26 | 33.67 | 31.08 | 28.49 | 15 |
| | | 7 | 20 | 40.29 | -3.65 | 88 | 36.64 | 32.99 | 29.34 | 25.69 | 5 |
| | | 14 | 20 | 40.22 | -3.95 | 79 | 36.27 | 32.31 | 28.36 | 24.41 | 5 |
| | | 21 | 23 | 39.87 | -3.81 | 83 | 36.06 | 32.25 | 28.43 | 24.62 | 2 |

* The regression parameters were calculated on the basis of the $log_{10}$ copy numbers per reaction.
** Level of Detection (LOD)95 was calculated based on effective volumes used for the PCR, whereas the other columns represent results based on quantitative Ct values.

#### (C) Result of Study-2- pre-shipment testing

A set of Inter-Laboratory Comparison Round-2 (ILC2) samples was tested at Moffett Proficiency Testing (PT) Laboratory prior to shipping as described in the methods section.

| Sample # | Description | Results |
|---|---|---|
| VM01 | Blank | ND |
| VM11 | Blank | ND |
| VM05 | Blank + FIPV | ND (for SARS-CoV-2); D (for FIPV) |
| VM10 | Blank + FIPV | ND (for SARS-CoV-2); D (for FIPV) |
| VM06 | 10 copies (B.1) /50 µL MTM | ND |
| VM09 | 10 copies (B.1) /50 µL MTM | ND |
| VM08 | 50 copies (B.1) /50 µL MTM | D |
| VM16 | 50 copies (B.1) /50 µL MTM | D |
| VM20 | 50 copies (B.1) /50 µL MTM | D |
| VM07 | 100 copies (B.1) /50 µL MTM | D |
| VM19 | 100 copies (B.1) /50 µL MTM | D |
| VM03 | 200 copies (B.1) /50 µL MTM | D |
| VM12 | 200 copies (B.1) /50 µL MTM | D |
| VM04 | 500 copies (B.1) /50 µL MTM | D |
| VM18 | 500 copies (B.1) /50 µL MTM | D |
| VM17 | 10,000 copies (B.1) /50 µL MTM | D |

| | | |
|---|---|---|
| VM15 | Alpha (B.1.1.7) 500 copies/50 µL MTM | D |
| VM13 | Alpha (B.1.1.7) 10,000 copies/50 µL MTM | D |
| VM14 | Beta (B.1.351) 500 copies/50 µL MTM | D |
| VM02 | Beta (B.1.351) 10,000 copies/50 µL MTM | D |

D: Determined by either N1 and/or N2 marker; ND: not detected by both N1 and N2 markers

FIPV: feline infectious peritonitis coronavirus

#### (D) **Results of Study-3- post-shipment testing**

A set of ICE2 samples was tested at Moffett PT Laboratory after shipping. Same procedures were performed as described for the Pre-shipment test.

| **Sample #** | **Description** | **Results** |
|---|---|---|
| VM01 | Blank | ND |
| VM11 | Blank | ND |
| VM05 | Blank + FIPV | ND (for SARS-CoV-2); D (for FIPV) |
| VM10 | Blank + FIPV | ND (for SARS-CoV-2); D (for FIPV) |
| VM06 | 10 copies (B.1) /50 µL MTM | ND |
| VM09 | 10 copies (B.1) /50 µL MTM | D |
| VM08 | 50 copies (B.1) /50 µL MTM | D |
| VM16 | 50 copies (B.1) /50 µL MTM | D |
| VM20 | 50 copies (B.1) /50 µL MTM | D |
| VM07 | 100 copies (B.1) /50 µL MTM | D |
| VM19 | 100 copies (B.1) /50 µL MTM | D |
| VM03 | 200 copies (B.1) /50 µL MTM | D |

| | | |
|---|---|---|
| VM12 | 200 copies (B.1) /50 µL MTM | D |
| VM04 | 500 copies (B.1) /50 µL MTM | D |
| VM18 | 500 copies (B.1) /50 µL MTM | D |
| VM17 | 10,000 copies (B.1) /50 µL MTM | D |
| VM15 | Alpha (B.1.1.7) 500 copies/50 µL MTM | D |
| VM13 | Alpha (B.1.1.7) 10,000 copies/50 µL MTM | D |
| VM14 | Beta (B.1.351) 500 copies/50 µL MTM | D |
| VM02 | Beta (B.1.351) 10,000 copies/50 µL MTM | D |

D: Determined by either N1 and/or N2 marker; ND: not detected by both N1 and N2 markers.

**Supplemental Table 2.** Ct values submitted by the analysts for the N1 and N2 markers

| Marker | Sample | Analyst Identification Number (AIN) | | | | | | | | | | | | | | | | | | | | | | | | | | | | | | | | | | | | | | | | | | | | | | | | | | | | | | |
|---|---|---|---|---|---|---|---|---|---|---|---|---|---|---|---|---|---|---|---|---|---|---|---|---|---|---|---|---|---|---|---|---|---|---|---|---|---|---|---|---|---|---|---|---|---|---|---|---|---|---|---|---|---|---|---|---|---|---|
| | | 240 | 241 | 242 | 243 | 244 | 245 | 246 | 247 | 248 | 249 | 250 | 251 | 252 | 253 | 254 | 255 | 256 | 257 | 258 | 259 | 260 | 261 | 262 | 263 | 264 | 265 | 266 | 267 | 268 | 269 | 270 | 271 | 272 | 273 | 274 | 275 | 276 | 277 | 278 | 279 | 280 | 281 | 282 | 283 | 284 | 285 | 286 | 287 | 288 | 289 | 290 | 291 | 292 | 293 | 294 | 295 | 296 |
| N1 | VM01 | | n | n | | n | n | n | | | n | | | | n | n | n | n | n | n | | | n | | n | | n | n | n | n | n | | n | | n | n | | n | n | n | n | n | n | n | n | n | n | | | | n | n | n | | n | | | n |
| | VM11 | | n | n | | n | n | n | | | n | | | | n | n | n | n | n | n | | | n | | n | | n | n | n | n | n | | n | | n | n | | n | n | n | n | n | n | n | n | n | n | | | | n | n | n | | n | | | n |
| | VM05 | | n | n | | n | n | n | | | n | | | | n | n | n | n | n | n | | | n | | n | | n | n | n | n | n | | n | | n | n | | n | n | n | n | n | n | n | n | n | n | | | | n | n | n | | n | | | n |
| | VM10 | | n | n | | n | n | n | | | n | | | | n | n | n | n | n | n | | | n | | n | | n | n | n | n | n | | n | | n | n | | n | n | n | n | n | n | n | n | n | n | | | | n | n | n | | n | | | n |
| | VM06 | | n | n | | 36 | n | n | | | 40 | | | | 40 | n | 39 | n | n | n | | | 37 | | n | | 39 | n | n | 38 | n | | n | | n | n | | n | 38 | n | 35 | n | 35 | n | n | 36 | n | | | | n | n | 38 | | n | | | n |
| | VM09 | | n | n | | n | n | 39 | | | 40 | | | | n | n | 38 | n | n | 40 | | | 37 | | n | | n | n | n | 38 | n | | 36 | | n | n | | n | n | n | n | n | 36 | n | n | 36 | n | | | | n | n | --- | | n | | | 37 |
| | VM08 | | n | n | | 35 | n | n | | | 38 | | | | n | 37 | 37 | 39 | 38 | 40 | | | 37 | | n | | 36 | 39 | n | 37 | 37 | | 33 | | n | n | | n | 37 | n | 36 | 38 | 33 | n | n | 35 | 40 | | | | n | 37 | n | | n | | | 35 |
| | VM16 | | n | n | | 36 | n | n | | | n | | | | 40 | n | 37 | 39 | 38 | 38 | | | 36 | | n | | 37 | 38 | n | 37 | 36 | | 35 | | n | n | | n | 37 | n | 36 | n | 35 | 42 | 38 | 34 | 38 | | | | n | 37 | 37 | | n | | | 35 |
| | VM20 | | 38 | n | | 36 | n | 39 | | | 39 | | | | n | 38 | 38 | 40 | 38 | 37 | | | 37 | | n | | 37 | 39 | 36 | n | n | | 34 | | n | n | | 34 | 36 | 36 | 34 | n | 34 | n | 42 | 36 | 40 | | | | 37 | 37 | n | | n | | | 35 |
| | VM07 | | 37 | 38 | | 33 | 38 | 39 | | | 38 | | | | 40 | 37 | 36 | 39 | 37 | 38 | | | 37 | | n | | 35 | 37 | n | 35 | 35 | | 33 | | 37 | 35 | | 34 | 35 | 35 | 33 | 39 | 32 | 38 | n | 34 | 37 | | | | 37 | 36 | n | | 37 | | | 35 |
| | VM19 | | 39 | n | | 34 | n | n | | | 40 | | | | n | 36 | 38 | 37 | 36 | 38 | | | 35 | | n | | 35 | 37 | 38 | 35 | 37 | | 33 | | n | n | | n | 36 | 34 | 33 | 38 | 33 | 37 | n | 35 | 37 | | | | 36 | 38 | n | | 37 | | | 34 |
| | VM03 | | 37 | 37 | | 32 | 38 | 38 | | | 37 | | | | 39 | 36 | 36 | 35 | 35 | 36 | | | 32 | | 35 | | 37 | 37 | 38 | 34 | 35 | | 32 | | 36 | n | | 35 | 35 | 36 | 32 | 38 | 31 | 35 | 40 | 33 | 38 | | | | 35 | 36 | 38 | | 37 | | | 33 |
| | VM12 | | 38 | 38 | | 32 | 37 | 38 | | | 37 | | | | n | 36 | 36 | 37 | 36 | 36 | | | 34 | | 35 | | 37 | 37 | 36 | 36 | 35 | | 31 | | n | 36 | | 34 | 35 | 35 | 32 | 37 | 32 | 37 | 36 | 33 | 36 | | | | 35 | 35 | 37 | | 37 | | | 33 |
| | VM04 | | 35 | 39 | | 31 | 37 | 37 | | | 35 | | | | 36 | 35 | 35 | 35 | 34 | 36 | | | 32 | | 35 | | 33 | 36 | 34 | 33 | 33 | | 31 | | 38 | 35 | | 34 | 34 | 34 | 30 | 36 | 30 | 34 | 39 | 32 | 35 | | | | 36 | 36 | 38 | | 37 | | | 31 |
| | VM18 | | 36 | n | | 32 | 37 | 38 | | | 36 | | | | 36 | 36 | 35 | 35 | 34 | 35 | | | 33 | | 34 | | 36 | 34 | 30 | 33 | 34 | | 31 | | n | 35 | | 33 | 33 | 35 | 31 | 37 | 30 | 34 | 41 | 32 | 35 | | | | 34 | 36 | 38 | | 37 | | | 32 |
| | VM17 | | 31 | 32 | | 27 | 32 | 33 | | | 31 | | | | 30 | 31 | 30 | 30 | 30 | 30 | | | 28 | | 30 | | 29 | 32 | 35 | 29 | 31 | | 26 | | 32 | 31 | | 29 | 30 | 32 | 27 | 32 | 26 | 31 | 33 | 27 | 31 | | | | 30 | 30 | 35 | | 33 | | | 27 |
| | VM15 | | 35 | 36 | | 32 | 36 | 37 | | | 36 | | | | 35 | 35 | 36 | 35 | 36 | 35 | | | 34 | | 34 | | 34 | 35 | 35 | 33 | 33 | | 31 | | 38 | 35 | | 33 | 34 | 34 | 33 | 36 | 31 | 35 | 37 | 32 | 35 | | | | 34 | 35 | 37 | | 39 | | | 33 |
| | VM13 | | 31 | 33 | | 27 | 32 | 33 | | | 32 | | | | 31 | 31 | 30 | 31 | 31 | 31 | | | 29 | | 30 | | 29 | 32 | 30 | 29 | 29 | | 26 | | 32 | 32 | | 29 | 30 | 31 | 28 | 32 | 26 | 30 | 33 | 28 | 31 | | | | 30 | 30 | 35 | | 35 | | | 28 |
| | VM14 | | 35 | 38 | | 32 | 38 | 37 | | | 36 | | | | n | 38 | 35 | 35 | 34 | 36 | | | 33 | | 35 | | 34 | 36 | 36 | 33 | 34 | | 31 | | 37 | 37 | | 33 | 35 | 34 | 31 | 39 | 30 | 34 | 36 | 32 | 36 | | | | 36 | 35 | 38 | | 36 | | | 32 |
| | VM02 | | 31 | 33 | | 28 | 32 | 33 | | | 32 | | | | 31 | 31 | 30 | 30 | 30 | 31 | | | 28 | | 30 | | 30 | 32 | 30 | 29 | 30 | | 27 | | 32 | 31 | | 30 | 30 | 31 | 27 | 32 | 26 | 31 | 33 | 27 | 31 | | | | 30 | 31 | 34 | | 33 | | | 28 |
| N2 | VM01 | | n | n | | n | n | n | | | | | | | n | n | n | n | n | n | | | n | | n | | n | n | n | n | n | | n | | n | n | | n | n | n | n | n | n | n | n | n | n | | | | n | n | n | | n | | | n |
| | VM11 | | n | n | | n | n | n | | | | | | | n | n | n | n | n | n | | | n | | n | | n | n | n | n | n | | n | | n | n | | n | n | n | n | n | n | n | n | n | n | | | | n | n | n | | n | | | n |
| | VM05 | | n | n | | n | n | n | | | | | | | n | n | n | n | n | n | | | n | | n | | n | n | 33 | n | n | | n | | n | n | | n | n | n | n | n | n | n | n | n | n | | | | n | n | n | | n | | | 36 |
| | VM10 | | n | n | | n | n | n | | | | | | | n | n | n | n | n | n | | | n | | n | | n | n | n | n | n | | n | | n | n | | n | n | n | n | n | n | n | n | n | n | | | | n | n | n | | n | | | n |
| | VM06 | | n | 38 | | 38 | n | 37 | | | | | | | n | n | 39 | n | n | n | | | 36 | | n | | 38 | n | 34 | 39 | n | | n | | n | n | | n | n | 38 | 37 | 40 | 36 | n | n | 39 | n | | | | n | 40 | n | | n | | | n |
| | VM09 | | n | n | | n | n | 40 | | | | | | | 36 | 40 | n | 40 | n | n | | | 37 | | n | | 38 | n | n | 38 | n | | 37 | | n | n | | n | 39 | n | n | n | n | n | 39 | 36 | n | | | | n | n | --- | | n | | | 38 |
| | VM08 | | 38 | n | | 35 | 40 | 38 | | | | | | | 38 | 40 | 37 | 39 | 38 | 38 | | | 35 | | 34 | | 36 | 40 | 41 | 37 | 33 | | 34 | | n | 41 | | n | 39 | n | 37 | 39 | 34 | n | 38 | 35 | 40 | | | | n | 37 | n | | 38 | | | 34 |
| | VM16 | | 37 | 39 | | 35 | 40 | n | | | | | | | 38 | 39 | 36 | 39 | 38 | n | | | 33 | | n | | 36 | 37 | 38 | 35 | 34 | | 36 | | 38 | n | | n | 39 | n | 37 | n | 36 | 42 | 38 | 35 | 40 | | | | 37 | 39 | n | | 38 | | | 35 |
| | VM20 | | 37 | n | | 35 | 40 | 40 | | | | | | | n | 40 | 37 | 39 | 38 | 38 | | | 36 | | n | | 36 | 41 | n | 36 | n | | 35 | | 38 | n | | n | 39 | 37 | 35 | n | 36 | n | n | 36 | 39 | | | | 38 | 38 | 37 | | n | | | 35 |
| | VM07 | | 36 | 39 | | 34 | 38 | 36 | | | | | | | 36 | 37 | 44 | 38 | 36 | 40 | | | 34 | | 36 | | 37 | 39 | 34 | 35 | 36 | | 34 | | 38 | 37 | | 34 | 36 | 39 | 35 | 40 | 33 | 45 | 38 | 34 | 39 | | | | 36 | 36 | n | | 38 | | | 34 |
| | VM19 | | 37 | 38 | | 34 | 39 | 35 | | | | | | | 37 | 38 | 38 | 38 | 36 | 37 | | | 34 | | 34 | | 36 | 40 | 38 | 35 | 34 | | 35 | | 38 | 39 | | 35 | 37 | 37 | 34 | 37 | 34 | 38 | 38 | 36 | 39 | | | | 36 | 37 | 38 | | 38 | | | 34 |
| | VM03 | | 34 | 37 | | 32 | 37 | 35 | | | | | | | 35 | 38 | 36 | 38 | 36 | 37 | | | 33 | | 33 | | 35 | 37 | 37 | 34 | 29 | | 32 | | 39 | 37 | | 37 | 37 | 38 | 34 | 37 | 33 | 36 | 37 | 34 | 38 | | | | 35 | 36 | 37 | | 35 | | | 33 |
| | VM12 | | 35 | 37 | | 33 | 38 | 35 | | | | | | | 37 | 39 | 36 | 37 | 36 | 37 | | | 33 | | 35 | | 34 | 38 | 37 | 34 | 18 | | 33 | | 40 | 36 | | n | 36 | 39 | 33 | 39 | 33 | 37 | 36 | 35 | 38 | | | | 35 | 36 | 37 | | 37 | | | 33 |
| | VM04 | | 34 | 37 | | 30 | 36 | 34 | | | | | | | 34 | 36 | 34 | 36 | 34 | 36 | | | 32 | | 33 | | 33 | 36 | 35 | 32 | 23 | | 32 | | 35 | 36 | | 35 | 35 | 38 | 32 | 35 | 32 | 34 | 36 | 32 | 37 | | | | 34 | 34 | 37 | | 36 | | | 31 |
| | VM18 | | 33 | 38 | | 31 | 36 | 33 | | | | | | | 35 | 37 | 35 | 36 | 34 | 37 | | | 32 | | 32 | | 33 | 36 | 30 | 33 | 32 | | 31 | | 35 | 37 | | 34 | 34 | 35 | 32 | 35 | 31 | 36 | 35 | 32 | 35 | | | | 34 | 35 | 37 | | 36 | | | 31 |
| | VM17 | | 29 | 32 | | 27 | 32 | 29 | | | | | | | 30 | 31 | 30 | 31 | 29 | 31 | | | 28 | | 28 | | 27 | 31 | 35 | 28 | 27 | | 27 | | 32 | 32 | | 29 | 30 | n | 28 | 31 | 27 | 32 | 30 | 27 | 31 | | | | 29 | 30 | 33 | | 32 | | | 27 |
| | VM15 | | 34 | 38 | | 32 | 36 | 34 | | | | | | | 34 | 38 | 41 | 37 | 37 | 36 | | | 33 | | 35 | | 34 | 36 | 36 | 33 | 30 | | 33 | | 37 | 37 | | 35 | 35 | 40 | 34 | 36 | 33 | 36 | 36 | 33 | 36 | | | | 35 | 36 | 38 | | 37 | | | 32 |
| | VM13 | | 30 | 33 | | 27 | 33 | 30 | | | | | | | 31 | 33 | 31 | 33 | 30 | 32 | | | 28 | | 29 | | 29 | 32 | 30 | 29 | 22 | | 28 | | 33 | 33 | | 30 | 31 | 33 | 29 | 32 | 28 | 31 | 32 | 28 | 32 | | | | 30 | 31 | 34 | | 34 | | | 28 |
| | VM14 | | 34 | 37 | | 31 | 36 | 34 | | | | | | | 35 | 39 | 36 | 36 | 34 | 37 | | | 32 | | 35 | | 32 | 36 | 32 | 33 | 28 | | 32 | | 37 | 38 | | 34 | 35 | 38 | 32 | 37 | 32 | 35 | 36 | 32 | 36 | | | | 34 | 35 | 38 | | 36 | | | 32 |
| | VM02 | | 29 | 32 | | 27 | 32 | 30 | | | | | | | 30 | 32 | 30 | 32 | 30 | 32 | | | 28 | | 29 | | 28 | 31 | 31 | 28 | 27 | | 27 | | 32 | 32 | | 30 | 30 | 32 | 28 | 32 | 27 | 31 | 32 | 27 | 31 | | | | 29 | 30 | 33 | | 32 | | | 28 |

Yellow color – false negative results (Ct was not reported or value above analyst-specific cut-off); pink color – false positive results (Ct value was reported and below analyst-specific cut-off); gray color – no results submitted for marker; n – no Ct value was reported; --- – no Ct value submitted for the particular sample.

**Supplemental Table 3. Ct values submitted by the analysts for the N and N3 markers**

| Marker | Sample | Analyst Identification Number (AIN) | | | | | | | | | | | | | | | | | | | | | | | | | | | |
|---|---|---|---|---|---|---|---|---|---|---|---|---|---|---|---|---|---|---|---|---|---|---|---|---|---|---|---|---|---|
| | | 240 | 241 | 242 | 243 | 244 | 245 | 246 | 247 | 248 | 249 | 250 | 251 | 252 | 253 | 254 | 255 | 256 | 257 | 258 | 259 | 260 | 261 | 262 | 263 | 264 | 265 | 266 | 267 |
| N | VM01 | n | | | 40 | | | | | n | n | n | n | | | | | | | | | n | | n | | n | | | |
| | VM11 | n | | | n | | | | | n | n | n | n | | | | | | | | | n | | --- | | n | | | |
| | VM05 | n | | | n | | | | | n | n | n | n | | | | | | | | | n | | n | | n | | | |
| | VM10 | n | | | n | | | | | n | n | n | n | | | | | | | | | n | | n | | n | | | |
| | VM06 | n | | | n | | | | | 39 | 39 | n | n | | | | | | | | | n | | 39 | | n | | | |
| | VM09 | n | | | n | | | | | 37 | n | n | n | | | | | | | | | n | | --- | | n | | | |
| | VM08 | 36 | | | 36 | | | | | 35 | 38 | 38 | n | | | | | | | | | n | | 38 | | 39 | | | |
| | VM16 | 35 | | | 37 | | | | | 35 | 39 | 37 | n | | | | | | | | | 39 | | 37 | | n | | | |
| | VM20 | n | | | 35 | | | | | 36 | n | n | n | | | | | | | | | n | | 37 | | n | | | |
| | VM07 | 38 | | | 34 | | | | | 32 | 37 | 37 | 34 | | | | | | | | | 37 | | 36 | | 35 | | | |
| | VM19 | 34 | | | 33 | | | | | 35 | 38 | n | 34 | | | | | | | | | 36 | | 37 | | n | | | |
| | VM03 | 34 | | | 33 | | | | | 34 | 36 | n | 31 | | | | | | | | | 36 | | 35 | | 35 | | | |
| | VM12 | 35 | | | 32 | | | | | 34 | 37 | n | 32 | | | | | | | | | 40 | | --- | | 35 | | | |
| | VM04 | 32 | | | 32 | | | | | 32 | 35 | 36 | 31 | | | | | | | | | 34 | | 33 | | 34 | | | |
| | VM18 | 33 | | | 31 | | | | | 32 | 37 | 35 | 32 | | | | | | | | | 34 | | 33 | | 33 | | | |
| | VM17 | 29 | | | 27 | | | | | 29 | 32 | 31 | 28 | | | | | | | | | 29 | | 29 | | 29 | | | |
| | VM15 | 33 | | | 32 | | | | | 34 | 37 | 36 | 30 | | | | | | | | | 35 | | 35 | | 33 | | | |
| | VM13 | 29 | | | 27 | | | | | 29 | 33 | 31 | 27 | | | | | | | | | 28 | | 30 | | 29 | | | |
| | VM14 | 34 | | | 32 | | | | | 33 | 37 | 35 | 32 | | | | | | | | | 33 | | 34 | | 34 | | | |
| | VM02 | 29 | | | 28 | | | | | 29 | 32 | 37 | 27 | | | | | | | | | 29 | | 29 | | --- | | | |
| N3 | VM01 | | | | | | | | | | | | | | | | n | | | | | | | | | | | | |
| | VM11 | | | | | | | | | | | | | | | | n | | | | | | | | | | | | |
| | VM05 | | | | | | | | | | | | | | | | n | | | | | | | | | | | | |
| | VM10 | | | | | | | | | | | | | | | | n | | | | | | | | | | | | |
| | VM06 | | | | | | | | | | | | | | | | 37 | | | | | | | | | | | | |
| | VM09 | | | | | | | | | | | | | | | | n | | | | | | | | | | | | |
| | VM08 | | | | | | | | | | | | | | | | 39 | | | | | | | | | | | | |
| | VM16 | | | | | | | | | | | | | | | | 36 | | | | | | | | | | | | |
| | VM20 | | | | | | | | | | | | | | | | 38 | | | | | | | | | | | | |
| | VM07 | | | | | | | | | | | | | | | | 37 | | | | | | | | | | | | |
| | VM19 | | | | | | | | | | | | | | | | 38 | | | | | | | | | | | | |
| | VM03 | | | | | | | | | | | | | | | | 36 | | | | | | | | | | | | |
| | VM12 | | | | | | | | | | | | | | | | 35 | | | | | | | | | | | | |
| | VM04 | | | | | | | | | | | | | | | | 34 | | | | | | | | | | | | |
| | VM18 | | | | | | | | | | | | | | | | 34 | | | | | | | | | | | | |
| | VM17 | | | | | | | | | | | | | | | | 30 | | | | | | | | | | | | |
| | VM15 | | | | | | | | | | | | | | | | 34 | | | | | | | | | | | | |
| | VM13 | | | | | | | | | | | | | | | | 31 | | | | | | | | | | | | |
| | VM14 | | | | | | | | | | | | | | | | 34 | | | | | | | | | | | | |
| | VM02 | | | | | | | | | | | | | | | | 30 | | | | | | | | | | | | |

| Marker | Sample | 268 | 269 | 270 | 271 | 272 | 273 | 274 | 275 | 276 | 277 | 278 | 279 | 280 | 281 | 282 | 283 | 284 | 285 | 286 | 287 | 288 | 289 | 290 | 291 | 292 | 293 | 294 | 295 | 296 |
|---|---|---|---|---|---|---|---|---|---|---|---|---|---|---|---|---|---|---|---|---|---|---|---|---|---|---|---|---|---|---|
| N | VM01 | | | | | n | | | n | | | | | | | | | | | | n | | | | | | | n | n | |
| | VM11 | | | | | n | | | n | | | | | | | | | | | | n | | | | | | | 39 | n | |
| | VM05 | | | | | n | | | n | | | | | | | | | | | | n | | | | | | | n | n | |
| | VM10 | | | | | n | | | n | | | | | | | | | | | | n | | | | | | | n | n | |
| | VM06 | | | | | n | | | 37 | | | | | | | | | | | | 37 | | | | | | | n | n | |
| | VM09 | | | | | n | | | 39 | | | | | | | | | | | | 36 | | | | | | | n | 36 | |
| | VM08 | | | | | 40 | | | 35 | | | | | | | | | | | | 34 | | | | | | | 36 | 35 | |
| | VM16 | | | | | 40 | | | 36 | | | | | | | | | | | | 34 | | | | | | | n | 37 | |
| | VM20 | | | | | n | | | 36 | | | | | | | | | | | | 36 | | | | | | | 38 | 39 | |
| | VM07 | | | | | n | | | 34 | | | | | | | | | | | | 33 | | | | | | | 39 | 36 | |
| | VM19 | | | | | 37 | | | 34 | | | | | | | | | | | | 34 | | | | | | | 38 | 35 | |
| | VM03 | | | | | 33 | | | 33 | | | | | | | | | | | | 33 | | | | | | | 35 | 34 | |
| | VM12 | | | | | 34 | | | 33 | | | | | | | | | | | | 33 | | | | | | | 35 | 34 | |
| | VM04 | | | | | 33 | | | 32 | | | | | | | | | | | | 31 | | | | | | | 34 | 32 | |
| | VM18 | | | | | 34 | | | 32 | | | | | | | | | | | | 31 | | | | | | | 34 | 33 | |
| | VM17 | | | | | 29 | | | 27 | | | | | | | | | | | | 27 | | | | | | | 29 | 28 | |
| | VM15 | | | | | 33 | | | 32 | | | | | | | | | | | | 32 | | | | | | | 35 | 33 | |
| | VM13 | | | | | 29 | | | 28 | | | | | | | | | | | | 28 | | | | | | | 31 | 29 | |
| | VM14 | | | | | 34 | | | 32 | | | | | | | | | | | | 32 | | | | | | | 34 | 33 | |
| | VM02 | | | | | 29 | | | 28 | | | | | | | | | | | | 27 | | | | | | | 30 | 28 | |
| N3 | VM01 | | | | | | | | | | | | | | | | | | | | | | | | | | | | | |
| | VM11 | | | | | | | | | | | | | | | | | | | | | | | | | | | | | |
| | VM05 | | | | | | | | | | | | | | | | | | | | | | | | | | | | | |
| | VM10 | | | | | | | | | | | | | | | | | | | | | | | | | | | | | |
| | VM06 | | | | | | | | | | | | | | | | | | | | | | | | | | | | | |
| | VM09 | | | | | | | | | | | | | | | | | | | | | | | | | | | | | |
| | VM08 | | | | | | | | | | | | | | | | | | | | | | | | | | | | | |
| | VM16 | | | | | | | | | | | | | | | | | | | | | | | | | | | | | |
| | VM20 | | | | | | | | | | | | | | | | | | | | | | | | | | | | | |
| | VM07 | | | | | | | | | | | | | | | | | | | | | | | | | | | | | |
| | VM19 | | | | | | | | | | | | | | | | | | | | | | | | | | | | | |
| | VM03 | | | | | | | | | | | | | | | | | | | | | | | | | | | | | |
| | VM12 | | | | | | | | | | | | | | | | | | | | | | | | | | | | | |
| | VM04 | | | | | | | | | | | | | | | | | | | | | | | | | | | | | |
| | VM18 | | | | | | | | | | | | | | | | | | | | | | | | | | | | | |
| | VM17 | | | | | | | | | | | | | | | | | | | | | | | | | | | | | |
| | VM15 | | | | | | | | | | | | | | | | | | | | | | | | | | | | | |
| | VM13 | | | | | | | | | | | | | | | | | | | | | | | | | | | | | |
| | VM14 | | | | | | | | | | | | | | | | | | | | | | | | | | | | | |
| | VM02 | | | | | | | | | | | | | | | | | | | | | | | | | | | | | |

Yellow color – false negative results (Ct was not reported or value above analyst-specific cut-off); pink color – false positive results (Ct value was reported and below analyst-specific cut-off); gray color – no results submitted for marker; n – no Ct value was reported; --- – no Ct value was submitted for the particular sample.

### Supplemental Table 4. Ct values submitted by the analysts for the E and RdRP markers

| Marker | Sample | Analyst Identification Number (AIN) | | | | | | | | | | | | | | | | | | | | | | | | | | | | | | | | | | | | | | | | | | | | | | | | | | | | | | |
|---|---|---|---|---|---|---|---|---|---|---|---|---|---|---|---|---|---|---|---|---|---|---|---|---|---|---|---|---|---|---|---|---|---|---|---|---|---|---|---|---|---|---|---|---|---|---|---|---|---|---|---|---|---|---|---|---|---|---|
| | | 240 | 241 | 242 | 243 | 244 | 245 | 246 | 247 | 248 | 249 | 250 | 251 | 252 | 253 | 254 | 255 | 256 | 257 | 258 | 259 | 260 | 261 | 262 | 263 | 264 | 265 | 266 | 267 | 268 | 269 | 270 | 271 | 272 | 273 | 274 | 275 | 276 | 277 | 278 | 279 | 280 | 281 | 282 | 283 | 284 | 285 | 286 | 287 | 288 | 289 | 290 | 291 | 292 | 293 | 294 | 295 | 296 |
| E | VM01 | | | | | | | | | | | | | n | | | | | | | n | | | | | | | | | | | n | | | | | | | | | | | | | | | | n | | n | | | | n | | | | |
| | VM11 | | | | | | | | | | | | | n | | | | | | | n | | | | | | | | | | | n | | | | | | | | | | | | | | | | n | | n | | | | n | | | | |
| | VM05 | | | | | | | | | | | | | n | | | | | | | n | | | | | | | | | | | n | | | | | | | | | | | | | | | | n | | n | | | | n | | | | |
| | VM10 | | | | | | | | | | | | | n | | | | | | | n | | | | | | | | | | | n | | | | | | | | | | | | | | | | n | | n | | | | n | | | | |
| | VM06 | | | | | | | | | | | | | n | | | | | | | 38 | | | | | | | | | | | n | | | | | | | | | | | | | | | | n | | 35 | | | | 35 | | | | |
| | VM09 | | | | | | | | | | | | | n | | | | | | | n | | | | | | | | | | | n | | | | | | | | | | | | | | | | n | | 35 | | | | 34 | | | | |
| | VM08 | | | | | | | | | | | | | 40 | | | | | | | 36 | | | | | | | | | | | 33 | | | | | | | | | | | | | | | | 35 | | 35 | | | | 34 | | | | |
| | VM16 | | | | | | | | | | | | | 37 | | | | | | | 36 | | | | | | | | | | | 34 | | | | | | | | | | | | | | | | 34 | | 34 | | | | 33 | | | | |
| | VM20 | | | | | | | | | | | | | 38 | | | | | | | 37 | | | | | | | | | | | 33 | | | | | | | | | | | | | | | | 35 | | 34 | | | | 34 | | | | |
| | VM07 | | | | | | | | | | | | | 36 | | | | | | | 35 | | | | | | | | | | | 32 | | | | | | | | | | | | | | | | 33 | | 34 | | | | 32 | | | | |
| | VM19 | | | | | | | | | | | | | 37 | | | | | | | 35 | | | | | | | | | | | 32 | | | | | | | | | | | | | | | | 34 | | 33 | | | | 32 | | | | |
| | VM03 | | | | | | | | | | | | | 35 | | | | | | | 34 | | | | | | | | | | | 31 | | | | | | | | | | | | | | | | 34 | | 32 | | | | 30 | | | | |
| | VM12 | | | | | | | | | | | | | 34 | | | | | | | 34 | | | | | | | | | | | 33 | | | | | | | | | | | | | | | | 33 | | 32 | | | | 31 | | | | |
| | VM04 | | | | | | | | | | | | | 33 | | | | | | | 33 | | | | | | | | | | | 30 | | | | | | | | | | | | | | | | 31 | | 31 | | | | 31 | | | | |
| | VM18 | | | | | | | | | | | | | 33 | | | | | | | 33 | | | | | | | | | | | 30 | | | | | | | | | | | | | | | | 31 | | 31 | | | | 31 | | | | |
| | VM17 | | | | | | | | | | | | | 29 | | | | | | | 29 | | | | | | | | | | | 25 | | | | | | | | | | | | | | | | 27 | | 27 | | | | 26 | | | | |
| | VM15 | | | | | | | | | | | | | 34 | | | | | | | 35 | | | | | | | | | | | 31 | | | | | | | | | | | | | | | | 32 | | 31 | | | | 31 | | | | |
| | VM13 | | | | | | | | | | | | | 29 | | | | | | | 31 | | | | | | | | | | | 26 | | | | | | | | | | | | | | | | 28 | | 27 | | | | 27 | | | | |
| | VM14 | | | | | | | | | | | | | 33 | | | | | | | 34 | | | | | | | | | | | 29 | | | | | | | | | | | | | | | | 33 | | 31 | | | | 30 | | | | |
| | VM02 | | | | | | | | | | | | | 29 | | | | | | | 29 | | | | | | | | | | | 25 | | | | | | | | | | | | | | | | 30 | | 26 | | | | 26 | | | | |
| RdRP | VM01 | | | | | | | | n | | | | | n | | | | | | | n | | | | | | | | | | | n | | | | | | | | | | | | | | | | | | n | | | | n | | | | |
| | VM11 | | | | | | | | n | | | | | n | | | | | | | n | | | | | | | | | | | n | | | | | | | | | | | | | | | | | | n | | | | n | | | | |
| | VM05 | | | | | | | | n | | | | | n | | | | | | | n | | | | | | | | | | | n | | | | | | | | | | | | | | | | | | n | | | | n | | | | |
| | VM10 | | | | | | | | n | | | | | n | | | | | | | n | | | | | | | | | | | n | | | | | | | | | | | | | | | | | | n | | | | n | | | | |
| | VM06 | | | | | | | | 41 | | | | | 39 | | | | | | | n | | | | | | | | | | | n | | | | | | | | | | | | | | | | | | 37 | | | | 36 | | | | |
| | VM09 | | | | | | | | n | | | | | 40 | | | | | | | n | | | | | | | | | | | n | | | | | | | | | | | | | | | | | | 37 | | | | n | | | | |
| | VM08 | | | | | | | | n | | | | | 38 | | | | | | | n | | | | | | | | | | | 36 | | | | | | | | | | | | | | | | | | 37 | | | | 35 | | | | |
| | VM16 | | | | | | | | n | | | | | 38 | | | | | | | 42 | | | | | | | | | | | n | | | | | | | | | | | | | | | | | | 37 | | | | 35 | | | | |
| | VM20 | | | | | | | | 37 | | | | | 39 | | | | | | | n | | | | | | | | | | | n | | | | | | | | | | | | | | | | | | 35 | | | | 35 | | | | |
| | VM07 | | | | | | | | 37 | | | | | 38 | | | | | | | n | | | | | | | | | | | 35 | | | | | | | | | | | | | | | | | | 36 | | | | 34 | | | | |
| | VM19 | | | | | | | | 38 | | | | | 37 | | | | | | | 41 | | | | | | | | | | | 36 | | | | | | | | | | | | | | | | | | 36 | | | | 35 | | | | |
| | VM03 | | | | | | | | 37 | | | | | 37 | | | | | | | 39 | | | | | | | | | | | 36 | | | | | | | | | | | | | | | | | | 33 | | | | 31 | | | | |
| | VM12 | | | | | | | | 39 | | | | | 36 | | | | | | | 39 | | | | | | | | | | | 41 | | | | | | | | | | | | | | | | | | 34 | | | | 33 | | | | |
| | VM04 | | | | | | | | 36 | | | | | 36 | | | | | | | 38 | | | | | | | | | | | 36 | | | | | | | | | | | | | | | | | | 32 | | | | 31 | | | | |
| | VM18 | | | | | | | | 35 | | | | | 36 | | | | | | | 39 | | | | | | | | | | | 35 | | | | | | | | | | | | | | | | | | 33 | | | | 31 | | | | |
| | VM17 | | | | | | | | 32 | | | | | 31 | | | | | | | 34 | | | | | | | | | | | 30 | | | | | | | | | | | | | | | | | | 29 | | | | 27 | | | | |
| | VM15 | | | | | | | | 35 | | | | | 36 | | | | | | | 38 | | | | | | | | | | | n | | | | | | | | | | | | | | | | | | 33 | | | | 33 | | | | |
| | VM13 | | | | | | | | 33 | | | | | 32 | | | | | | | 34 | | | | | | | | | | | 31 | | | | | | | | | | | | | | | | | | 29 | | | | 28 | | | | |
| | VM14 | | | | | | | | 36 | | | | | 35 | | | | | | | 38 | | | | | | | | | | | 36 | | | | | | | | | | | | | | | | | | 33 | | | | 32 | | | | |
| | VM02 | | | | | | | | 33 | | | | | 31 | | | | | | | 34 | | | | | | | | | | | 30 | | | | | | | | | | | | | | | | | | 28 | | | | 27 | | | | |

Yellow color – false negative results (Ct was not reported or value above analyst-specific cut-off); gray color – no results submitted for marker; n – no Ct value was reported; --- – no Ct value was submitted for the particular sample.

### Supplemental Table 5. Ct values submitted by the analysts for the ORF1ab and S markers.

| Marker | Sample | Analyst Identification Number (AIN) | | | | | | | | | | | | | | | | | | | | | | | | | | | | | | | | | | | | | | | | | | | | | | | | | | | | | | |
|---|---|---|---|---|---|---|---|---|---|---|---|---|---|---|---|---|---|---|---|---|---|---|---|---|---|---|---|---|---|---|---|---|---|---|---|---|---|---|---|---|---|---|---|---|---|---|---|---|---|---|---|---|---|---|---|---|---|---|
| | | 240 | 241 | 242 | 243 | 244 | 245 | 246 | 247 | 248 | 249 | 250 | 251 | 252 | 253 | 254 | 255 | 256 | 257 | 258 | 259 | 260 | 261 | 262 | 263 | 264 | 265 | 266 | 267 | 268 | 269 | 270 | 271 | 272 | 273 | 274 | 275 | 276 | 277 | 278 | 279 | 280 | 281 | 282 | 283 | 284 | 285 | 286 | 287 | 288 | 289 | 290 | 291 | 292 | 293 | 294 | 295 | 296 |
| ORF1ab | VM01 | n | | | 39 | | | | | | | | n | | | | | | | | | n | | | | n | | | | | | | | n | | | | | | | | | | | | | | n | | | | | | | | | | |
| | VM11 | n | | | n | | | | | | | | n | | | | | | | | | n | | | | n | | | | | | | | n | | | | | | | | | | | | | | n | | | | | | | | | | |
| | VM05 | n | | | n | | | | | | | | n | | | | | | | | | n | | | | n | | | | | | | | n | | | | | | | | | | | | | | 39 | | | | | | | | | | |
| | VM10 | n | | | n | | | | | | | | n | | | | | | | | | n | | | | n | | | | | | | | n | | | | | | | | | | | | | | 40 | | | | | | | | | | |
| | VM06 | 38 | | | 35 | | | | | | | | 35 | | | | | | | | | n | | | | n | | | | | | | | n | | | | | | | | | | | | | | 44 | | | | | | | | | | |
| | VM09 | 37 | | | n | | | | | | | | n | | | | | | | | | n | | | | n | | | | | | | | n | | | | | | | | | | | | | | n | | | | | | | | | | |
| | VM08 | 34 | | | 33 | | | | | | | | 33 | | | | | | | | | n | | | | 40 | | | | | | | | 35 | | | | | | | | | | | | | | 40 | | | | | | | | | | |
| | VM16 | 34 | | | 33 | | | | | | | | 32 | | | | | | | | | n | | | | n | | | | | | | | 36 | | | | | | | | | | | | | | 43 | | | | | | | | | | |
| | VM20 | 36 | | | 33 | | | | | | | | 34 | | | | | | | | | 39 | | | | 37 | | | | | | | | 37 | | | | | | | | | | | | | | 42 | | | | | | | | | | |
| | VM07 | 33 | | | 32 | | | | | | | | 33 | | | | | | | | | 37 | | | | 36 | | | | | | | | 35 | | | | | | | | | | | | | | 40 | | | | | | | | | | |
| | VM19 | 34 | | | 32 | | | | | | | | 34 | | | | | | | | | 38 | | | | 35 | | | | | | | | 36 | | | | | | | | | | | | | | 40 | | | | | | | | | | |
| | VM03 | 33 | | | 31 | | | | | | | | 29 | | | | | | | | | 36 | | | | 35 | | | | | | | | 34 | | | | | | | | | | | | | | 43 | | | | | | | | | | |
| | VM12 | 33 | | | 31 | | | | | | | | 31 | | | | | | | | | 38 | | | | 33 | | | | | | | | 34 | | | | | | | | | | | | | | 38 | | | | | | | | | | |
| | VM04 | 31 | | | 30 | | | | | | | | 31 | | | | | | | | | 33 | | | | 32 | | | | | | | | 33 | | | | | | | | | | | | | | 37 | | | | | | | | | | |
| | VM18 | 31 | | | 30 | | | | | | | | 32 | | | | | | | | | 32 | | | | 32 | | | | | | | | 33 | | | | | | | | | | | | | | 37 | | | | | | | | | | |
| | VM17 | 27 | | | 26 | | | | | | | | 27 | | | | | | | | | 28 | | | | 28 | | | | | | | | 29 | | | | | | | | | | | | | | 33 | | | | | | | | | | |
| | VM15 | 33 | | | 31 | | | | | | | | 29 | | | | | | | | | 34 | | | | 33 | | | | | | | | 34 | | | | | | | | | | | | | | 37 | | | | | | | | | | |
| | VM13 | 28 | | | 27 | | | | | | | | 27 | | | | | | | | | 29 | | | | 29 | | | | | | | | 30 | | | | | | | | | | | | | | 33 | | | | | | | | | | |
| | VM14 | 31 | | | 31 | | | | | | | | 31 | | | | | | | | | 33 | | | | 32 | | | | | | | | 33 | | | | | | | | | | | | | | 38 | | | | | | | | | | |
| | VM02 | 27 | | | 26 | | | | | | | | 26 | | | | | | | | | 28 | | | | --- | | | | | | | | 28 | | | | | | | | | | | | | | 33 | | | | | | | | | | |
| S | VM01 | n | n | | n | | | | | | | | n | | | | | | | | | n | | | | n | | | | | | | | n | | | | | | | | | | | | | | | | | | | | | | | | |
| | VM11 | n | n | | n | | | | | | | | n | | | | | | | | | n | | | | n | | | | | | | | n | | | | | | | | | | | | | | | | | | | | | | | | |
| | VM05 | n | n | | n | | | | | | | | n | | | | | | | | | n | | | | n | | | | | | | | n | | | | | | | | | | | | | | | | | | | | | | | | |
| | VM10 | n | n | | n | | | | | | | | n | | | | | | | | | n | | | | n | | | | | | | | n | | | | | | | | | | | | | | | | | | | | | | | | |
| | VM06 | 39 | n | | 39 | | | | | | | | n | | | | | | | | | n | | | | n | | | | | | | | n | | | | | | | | | | | | | | | | | | | | | | | | |
| | VM09 | n | n | | n | | | | | | | | n | | | | | | | | | n | | | | n | | | | | | | | n | | | | | | | | | | | | | | | | | | | | | | | | |
| | VM08 | 35 | 38 | | 34 | | | | | | | | n | | | | | | | | | 37 | | | | n | | | | | | | | 39 | | | | | | | | | | | | | | | | | | | | | | | | |
| | VM16 | 35 | n | | 34 | | | | | | | | 36 | | | | | | | | | n | | | | n | | | | | | | | 39 | | | | | | | | | | | | | | | | | | | | | | | | |
| | VM20 | 35 | 38 | | 35 | | | | | | | | n | | | | | | | | | n | | | | n | | | | | | | | n | | | | | | | | | | | | | | | | | | | | | | | | |
| | VM07 | 36 | n | | 33 | | | | | | | | n | | | | | | | | | 39 | | | | 34 | | | | | | | | 34 | | | | | | | | | | | | | | | | | | | | | | | | |
| | VM19 | 34 | n | | 33 | | | | | | | | 37 | | | | | | | | | 37 | | | | 38 | | | | | | | | 35 | | | | | | | | | | | | | | | | | | | | | | | | |
| | VM03 | 33 | 37 | | 32 | | | | | | | | 24 | | | | | | | | | 35 | | | | 33 | | | | | | | | 33 | | | | | | | | | | | | | | | | | | | | | | | | |
| | VM12 | 33 | 37 | | 32 | | | | | | | | 36 | | | | | | | | | 33 | | | | 35 | | | | | | | | 33 | | | | | | | | | | | | | | | | | | | | | | | | |
| | VM04 | 32 | 36 | | 31 | | | | | | | | 32 | | | | | | | | | 32 | | | | 32 | | | | | | | | 32 | | | | | | | | | | | | | | | | | | | | | | | | |
| | VM18 | 32 | 37 | | 30 | | | | | | | | 31 | | | | | | | | | 32 | | | | 31 | | | | | | | | 32 | | | | | | | | | | | | | | | | | | | | | | | | |
| | VM17 | 27 | 31 | | 26 | | | | | | | | 26 | | | | | | | | | 27 | | | | 27 | | | | | | | | 27 | | | | | | | | | | | | | | | | | | | | | | | | |
| | VM15 | n | 36 | | n | | | | | | | | n | | | | | | | | | n | | | | n | | | | | | | | n | | | | | | | | | | | | | | | | | | | | | | | | |
| | VM13 | n | 33 | | n | | | | | | | | n | | | | | | | | | n | | | | n | | | | | | | | n | | | | | | | | | | | | | | | | | | | | | | | | |
| | VM14 | 33 | 35 | | 31 | | | | | | | | 32 | | | | | | | | | 31 | | | | 31 | | | | | | | | 32 | | | | | | | | | | | | | | | | | | | | | | | | |
| | VM02 | 28 | 30 | | 26 | | | | | | | | 23 | | | | | | | | | 27 | | | | --- | | | | | | | | 27 | | | | | | | | | | | | | | | | | | | | | | | | |

Yellow color – false negative results (Ct was not reported or value above analyst-specific cut-off); pink color – false positive results (Ct value was reported and below analyst-specific cut-off); gray color – no results submitted for marker; n – no Ct value was reported; --- – no Ct value was submitted for the particular sample.

### Supplemental Table 6. Ct values submitted by the analysts for the other markers 1 and 2.

| Marker | Sample | Analyst Identification Number (AIN) | | | | | | | | | | | | | | | | | | | | | | | | | | | | | | | | | | | | | | | | | | | | | | | | | | | | | | |
|---|---|---|---|---|---|---|---|---|---|---|---|---|---|---|---|---|---|---|---|---|---|---|---|---|---|---|---|---|---|---|---|---|---|---|---|---|---|---|---|---|---|---|---|---|---|---|---|---|---|---|---|---|---|---|---|---|---|---|
| | | 240 | 241 | 242 | 243 | 244 | 245 | 246 | 247 | 248 | 249 | 250 | 251 | 252 | 253 | 254 | 255 | 256 | 257 | 258 | 259* | 260 | 261 | 262 | 263 | 264 | 265 | 266 | 267 | 268 | 269 | 270* | 271 | 272 | 273 | 274 | 275 | 276 | 277 | 278 | 279 | 280 | 281 | 282 | 283 | 284 | 285 | 286 | 287 | 288 | 289 | 290 | 291** | 292 | 293** | 294 | 295 | 296 |
| Other1 | VM01 | | | | | | | | | | | | | | | | | | | | n | | | | | | | | | | | | | | | | | | | | | | | | | | | | | | | | n | | n | | | |
| | VM11 | | | | | | | | | | | | | | | | | | | | n | | | | | | | | | | | | | | | | | | | | | | | | | | | | | | | | n | | n | | | |
| | VM05 | | | | | | | | | | | | | | | | | | | | n | | | | | | | | | | | | | | | | | | | | | | | | | | | | | | | | n | | n | | | |
| | VM10 | | | | | | | | | | | | | | | | | | | | n | | | | | | | | | | | | | | | | | | | | | | | | | | | | | | | | n | | n | | | |
| | VM06 | | | | | | | | | | | | | | | | | | | | n | | | | | | | | | | | | | | | | | | | | | | | | | | | | | | | | n | | n | | | |
| | VM09 | | | | | | | | | | | | | | | | | | | | n | | | | | | | | | | | | | | | | | | | | | | | | | | | | | | | | --- | | n | | | |
| | VM08 | | | | | | | | | | | | | | | | | | | | 40 | | | | | | | | | | | | | | | | | | | | | | | | | | | | | | | | n | | n | | | |
| | VM16 | | | | | | | | | | | | | | | | | | | | 39 | | | | | | | | | | | | | | | | | | | | | | | | | | | | | | | | 40 | | 40 | | | |
| | VM20 | | | | | | | | | | | | | | | | | | | | n | | | | | | | | | | | | | | | | | | | | | | | | | | | | | | | | 40 | | n | | | |
| | VM07 | | | | | | | | | | | | | | | | | | | | 39 | | | | | | | | | | | | | | | | | | | | | | | | | | | | | | | | 40 | | 40 | | | |
| | VM19 | | | | | | | | | | | | | | | | | | | | 39 | | | | | | | | | | | | | | | | | | | | | | | | | | | | | | | | 40 | | 40 | | | |
| | VM03 | | | | | | | | | | | | | | | | | | | | 38 | | | | | | | | | | | | | | | | | | | | | | | | | | | | | | | | 40 | | 40 | | | |
| | VM12 | | | | | | | | | | | | | | | | | | | | 38 | | | | | | | | | | | | | | | | | | | | | | | | | | | | | | | | 40 | | 40 | | | |
| | VM04 | | | | | | | | | | | | | | | | | | | | 36 | | | | | | | | | | | | | | | | | | | | | | | | | | | | | | | | 40 | | 39 | | | |
| | VM18 | | | | | | | | | | | | | | | | | | | | 37 | | | | | | | | | | | | | | | | | | | | | | | | | | | | | | | | 39 | | 37 | | | |
| | VM17 | | | | | | | | | | | | | | | | | | | | 33 | | | | | | | | | | | | | | | | | | | | | | | | | | | | | | | | 33 | | 31 | | | |
| | VM15 | | | | | | | | | | | | | | | | | | | | 38 | | | | | | | | | | | | | | | | | | | | | | | | | | | | | | | | n | | 37 | | | |
| | VM13 | | | | | | | | | | | | | | | | | | | | 33 | | | | | | | | | | | | | | | | | | | | | | | | | | | | | | | | 33 | | 32 | | | |
| | VM14 | | | | | | | | | | | | | | | | | | | | 37 | | | | | | | | | | | | | | | | | | | | | | | | | | | | | | | | 39 | | 40 | | | |
| | VM02 | | | | | | | | | | | | | | | | | | | | 34 | | | | | | | | | | | | | | | | | | | | | | | | | | | | | | | | 32 | | 31 | | | |
| Other2 | VM01 | | | | | | | | | | | | | | | | | | | | | | | | | | | | | | | n | | | | | | | | | | | | | | | | | | | | | n | | n | | | |
| | VM11 | | | | | | | | | | | | | | | | | | | | | | | | | | | | | | | n | | | | | | | | | | | | | | | | | | | | | n | | n | | | |
| | VM05 | | | | | | | | | | | | | | | | | | | | | | | | | | | | | | | n | | | | | | | | | | | | | | | | | | | | | n | | n | | | |
| | VM10 | | | | | | | | | | | | | | | | | | | | | | | | | | | | | | | n | | | | | | | | | | | | | | | | | | | | | n | | n | | | |
| | VM06 | | | | | | | | | | | | | | | | | | | | | | | | | | | | | | | n | | | | | | | | | | | | | | | | | | | | | n | | n | | | |
| | VM09 | | | | | | | | | | | | | | | | | | | | | | | | | | | | | | | n | | | | | | | | | | | | | | | | | | | | | --- | | n | | | |
| | VM08 | | | | | | | | | | | | | | | | | | | | | | | | | | | | | | | 36 | | | | | | | | | | | | | | | | | | | | | n | | n | | | |
| | VM16 | | | | | | | | | | | | | | | | | | | | | | | | | | | | | | | n | | | | | | | | | | | | | | | | | | | | | n | | n | | | |
| | VM20 | | | | | | | | | | | | | | | | | | | | | | | | | | | | | | | 36 | | | | | | | | | | | | | | | | | | | | | n | | n | | | |
| | VM07 | | | | | | | | | | | | | | | | | | | | | | | | | | | | | | | 42 | | | | | | | | | | | | | | | | | | | | | n | | n | | | |
| | VM19 | | | | | | | | | | | | | | | | | | | | | | | | | | | | | | | 36 | | | | | | | | | | | | | | | | | | | | | n | | n | | | |
| | VM03 | | | | | | | | | | | | | | | | | | | | | | | | | | | | | | | 34 | | | | | | | | | | | | | | | | | | | | | n | | 36 | | | |
| | VM12 | | | | | | | | | | | | | | | | | | | | | | | | | | | | | | | 35 | | | | | | | | | | | | | | | | | | | | | n | | n | | | |
| | VM04 | | | | | | | | | | | | | | | | | | | | | | | | | | | | | | | 34 | | | | | | | | | | | | | | | | | | | | | 36 | | 35 | | | |
| | VM18 | | | | | | | | | | | | | | | | | | | | | | | | | | | | | | | 35 | | | | | | | | | | | | | | | | | | | | | 37 | | 35 | | | |
| | VM17 | | | | | | | | | | | | | | | | | | | | | | | | | | | | | | | 30 | | | | | | | | | | | | | | | | | | | | | 35 | | 32 | | | |
| | VM15 | | | | | | | | | | | | | | | | | | | | | | | | | | | | | | | 34 | | | | | | | | | | | | | | | | | | | | | n | | n | | | |
| | VM13 | | | | | | | | | | | | | | | | | | | | | | | | | | | | | | | 30 | | | | | | | | | | | | | | | | | | | | | 35 | | 34 | | | |
| | VM14 | | | | | | | | | | | | | | | | | | | | | | | | | | | | | | | 34 | | | | | | | | | | | | | | | | | | | | | n | | 35 | | | |
| | VM02 | | | | | | | | | | | | | | | | | | | | | | | | | | | | | | | 30 | | | | | | | | | | | | | | | | | | | | | 34 | | 33 | | | |

Yellow color – false negative results (Ct was not reported or value above analyst-specific cut-off); grey color – no results submitted for marker; n – no Ct value was reported; --- – no Ct value was submitted for the particular sample.

* analysts 259 and 270: RdRP discriminatory assay; ** analysts 291 and 293: other marker 1 = SARS-Cov-2 Nucleocapsid; other marker 2 = SARS-Cov-2 7b regulatory gene.
